# Saliva TwoStep for rapid detection of asymptomatic SARS-CoV-2 carriers

**DOI:** 10.1101/2020.07.16.20150250

**Authors:** Qing Yang, Nicholas R. Meyerson, Stephen K. Clark, Camille L. Paige, Will T. Fattor, Alison R. Gilchrist, Arturo Barbachano-Guerrero, Benjamin G. Healy, Emma R. Worden-Sapper, Sharon S. Wu, Denise Muhlrad, Carolyn J. Decker, Tassa K. Saldi, Erika Lasda, Patrick K. Gonzales, Morgan R. Fink, Kimngan L. Tat, Cole R. Hager, Jack C. Davis, Christopher D. Ozeroff, Gloria R. Brisson, Matthew B. McQueen, Leslie Leinwand, Roy Parker, Sara L. Sawyer

**Affiliations:** BioFrontiers Institute, University of Colorado Boulder, Boulder, Colorado, 80303; Department of Molecular, Cellular, and Developmental Biology, University of Colorado Boulder, Boulder, Colorado, 80303; Wardenburg Health Center, University of Colorado Boulder, Boulder, Colorado, 80303; Department of Integrative Physiology, University of Colorado Boulder, Boulder, Colorado, 80303; Department of Biochemistry, University of Colorado Boulder, Boulder, Colorado, 80303; Howard Hughes Medical Institute, University of Colorado Boulder, Boulder, Colorado, 80303; Department of Mechanical Engineering, University of Colorado Boulder, Boulder, Colorado, 80303; Interdisciplinary Quantitative Biology Graduate Program, University of Colorado Boulder, Boulder, Colorado, 80303; Darwin Biosciences Inc., Boulder, Colorado, 80303, USA

## Abstract

Here, we develop a simple molecular test for SARS-CoV-2 in saliva based on reverse transcription loop-mediated isothermal amplification (RT-LAMP). The test has two steps: 1) heat saliva with a stabilization solution, and 2) detect virus by incubating with a primer/enzyme mix. After incubation, saliva samples containing the SARS-CoV-2 genome turn bright yellow. Because this test is pH dependent, it can react falsely to some naturally acidic saliva samples. We report unique saliva stabilization protocols that rendered 295 healthy saliva samples compatible with the test, producing zero false positives. We also evaluated the test on 278 saliva samples from individuals who were infected with SARS-CoV-2 but had no symptoms at the time of saliva collection, and from 54 matched pairs of saliva and anterior nasal samples from infected individuals. The Saliva TwoStep test described herein identified infections with 94% sensitivity and >99% specificity in individuals with sub-clinical (asymptomatic or pre-symptomatic) infections.

## Introduction

Disease screening is one of the most basic and powerful tools in the public health arsenal. Screening tests identify unknown illness in apparently healthy or asymptomatic individuals. In the case of dangerous pathogens, screening tests serve to direct potential carriers of the pathogen into the healthcare system for confirmatory testing, and to alert them that they could possibly infect others while they await confirmatory results. If dangerous pathogens are spreading at high rates, individuals will need to be screened frequently. As such, screening tests should operate with minimal requirements for laboratory equipment and labor, such that they are community-deployable and don’t burden the critical pipelines for diagnostics. In the current SARS-CoV-2 pandemic, body temperature is a ubiquitous screening test being used on apparently-healthy people around the world. However, using elevated body temperature as a sign of SARS-CoV-2 infection lacks specificity for this particular pathogen and sensitivity in identifying asymptomatic carriers (Wright and Mackowiak, 2020). To help fill in the need for more reliable screening tests, here we present a simple and portable assay that detects the SARS-CoV-2 genome in saliva with specificity and sensitivity, even in samples from individuals with no symptoms at the time of saliva collection.

LAMP (loop-mediated isothermal amplification) is a simple nucleic acid diagnostic concept that has existed for more than 20 years (Notomi et al., 2000). It has been used in diverse and even remote settings to test samples for the presence of viral nucleic acids (Brewster et al., 2018; Chotiwan et al., 2017). LAMP utilizes loop forming primers and strand-displacement polymerases to achieve isothermal amplification of a target nucleic acid template, and therefore does not require a thermal cycler. LAMP assays can be performed anywhere because they simply require pipettors and a heating source (e.g. water baths or heat blocks) as equipment (Brewster et al., 2018). LAMP assays offer robust amplification of target material and can produce on the order of 10^9^ copies of the target in an hour-long reaction (Notomi et al., 2000). Successful amplification in LAMP reactions can be directly visualized by simply looking at the reaction tube, where the reaction mix changes color upon successful target amplification. These colorimetric changes can be triggered by pH indicator dyes or metal ion indicators, which change color when successful target amplification changes the chemistry within the reaction tube (Kellner et al., 2020; Notomi et al., 2000; Tanner et al., 2015). If more sophisticated visualization equipment is available, other indicators can used. Intercalating fluorescent DNA dyes or quenched fluorescent probes can be used which emit fluorescent signal over time during amplification (Hardinge and Murray, 2019; Seyrig et al., 2015). Alternately, real-time measurements of turbidity in the tube can be used to measure changes in turbidity resulting from magnesium pyrophosphate formation as amplification proceeds (Mori et al., 2001). RT-LAMP (reverse transcription - loop-mediated isothermal amplification), where a reverse transcription step is added upstream of the LAMP reaction, adapts all of these protocols for detection of RNA. RT-LAMP with a simple visual color change that occurs in sample tubes containing SARS-CoV-2 could be well suited as a rapid and deployable community-based screening test (Khan et al., 2020).

Recent studies have shown that saliva has high diagnostic value for SARS-CoV-2 (Butler-Laporte et al., 2021; Silva et al., 2021; Vogels et al., 2020; Wyllie et al., 2020; Yokota et al., 2020). Compared to nasopharyngeal swabs, saliva samples harbor similar levels of viral load while being easier to obtain via self-collection. Several groups have developed RT-LAMP tests to detect SARS-CoV-2 in saliva samples (Bhadra et al., 2020; Flynn et al., n.d.; Lalli et al., 2020; Lamb et al., 2020; Nagura-Ikeda et al., 2020; Rabe and Cepko, 2020; Taki et al., 2020; Yokota et al., 2020). However, due to pH variability between saliva samples, RT-LAMP often has a high rate of false positives when used with the common pH-dependent dye phenol red (Bhadra et al., 2020; Hardinge and Murray, 2019). In RT-LAMP reactions containing phenol red, reactions start as pink/red but turn strongly yellow at pH 6.8 and below. When RT-LAMP amplifies a target, hydrogen ions are released during dNTP incorporation. This causes a drop in pH within the tube to pH 6.0 – 6.5, triggering the color change to yellow (Tanner et al., 2015). Human saliva naturally varies in pH between 6.8 and 7.4 (Cameron et al., 2015), posing a significant problem in this pH-dependent assay. In fact, we find about 7% of human saliva samples are naturally acidic enough to immediately trigger phenol red-containing reactions to change to yellow without any target amplification (**Figure 1A Left**). If acidic samples are not anticipated and managed, colorimetric RT-LAMP has the potential to produce a high false-positive rate.

**Figure 1:**
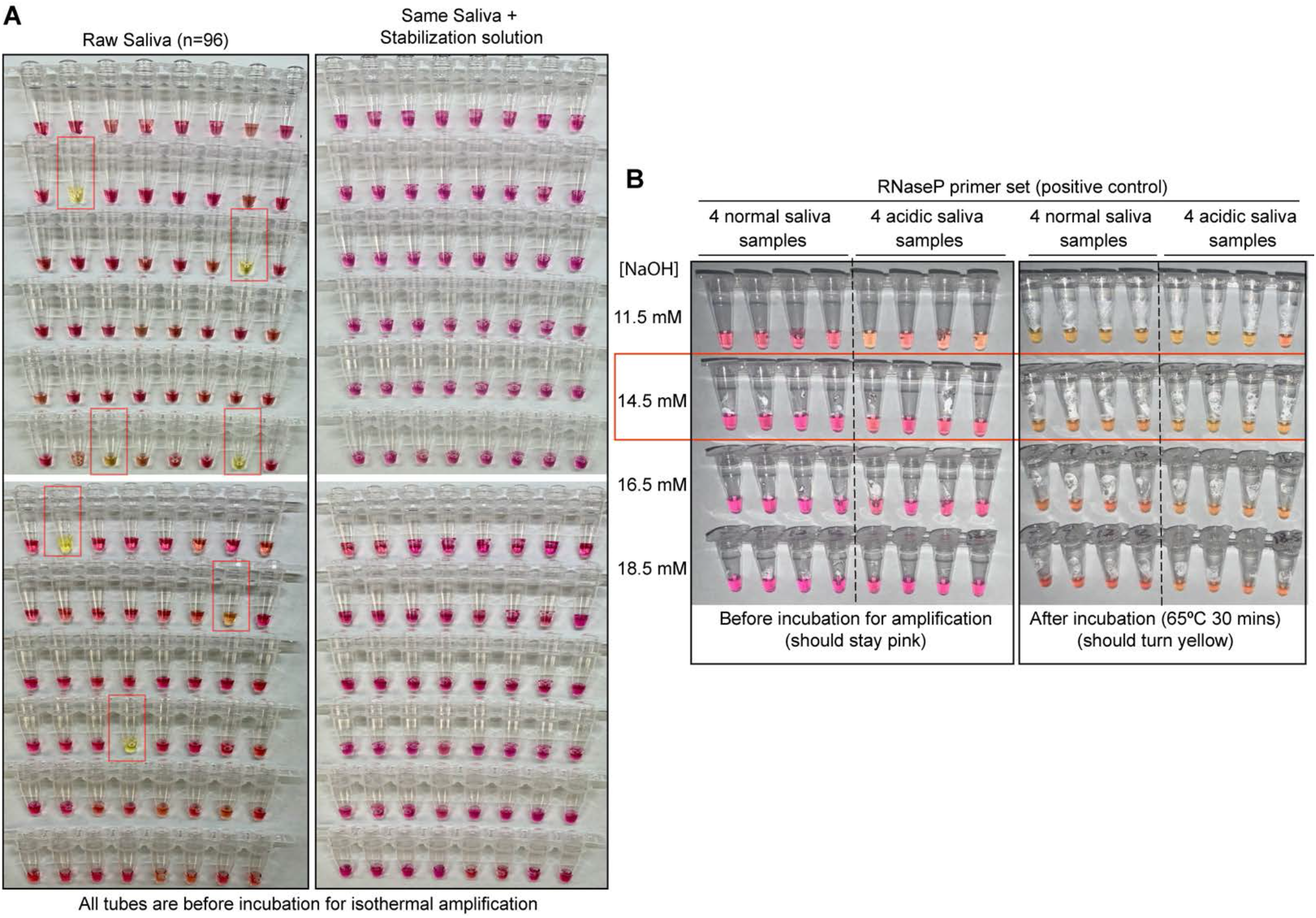
Optimized strategy for controlling natural variability in saliva pH. **A)** Here, saliva samples from 96 different individuals are analyzed for the prevalence of natural acidity extreme enough to trigger the pink-to-yellow color change of phenol red even before isothermal amplification. Each saliva sample was combined 1:1 with water (left) or 2X saliva stabilization solution (right; methods) and heated at 95°C for 10 minutes to liberate RNA from virions. 2 µL of each was then added to 18 µL RT-LAMP reaction mix (2X Colorimetric RT-LAMP Master Mix, RNase P primers, nuclease-free water). The pictures show tubes immediately after samples and master mix are combined, before any incubation steps are undertaken to commence isothermal amplification. With raw saliva, 7 out of 96 tubes turned yellow at this step (highlighted in red boxes). These are false-positives, because no amplification reaction has occurred. None of these 96 saliva samples mixed with saliva stabilization solution turned the reaction tube prematurely yellow. **B)** Here, we show the method that we had used to identify the ideal pH of the saliva stabilization solution used in panel A and throughout this paper. We chose 4 normal and 4 acidic saliva samples and mixed each 1:1 with 2X saliva stabilization solution containing NaOH at various concentrations (final molarity of NaOH after mixing shown). Samples were then heated at 95°C for 10 minutes and combined with RT-LAMP reaction mix and control primers recognizing the human RNase P transcript. Before incubation, all tubes should be pink, and after incubation all tubes should be yellow. Based on this, the red box indicates the final optimal NaOH concentration chosen.

Here, we combine the simplicity of RT-LAMP and the non-invasive nature of saliva to develop an effective screening test for SARS-CoV-2. This test does not require RNA purification but rather works directly with human saliva. We optimized a saliva stabilization solution that 1) neutralizes the variability of human saliva and essentially eliminates false positives, 2) lowers the viscosity of saliva, and 3) stabilizes RNA for analysis in the test. We validated the RT-LAMP test using a large cohort of saliva and matched nasal swab specimens collected from our local university population, comparing the test to two other quantitative RT-PCR-based SARS-CoV-2 tests (one nasal test and one saliva test). We found our optimized RT-LAMP procedure performs consistently with high specificity and sensitivity, even though our samples were largely from individuals who had no reported symptoms at the time of sample collection. Based on our experience performing screening on our university campus and elsewhere, we provide in the supplement extensive operational details and recommendations for successful community deployment of this SARS-CoV-2 screening test.

## Results

### Optimized universal saliva stabilization conditions for RT-LAMP

To deal with the variability in pH of human saliva, we optimized a basic saliva stabilization solution by titrating in various concentrations of sodium hydroxide (NaOH). We performed this optimization using a control RT-LAMP primer set, “RNaseP,” which amplifies the mRNA transcript produced from the human *POP7* gene (primer set developed previously (Curtis et al., 2018)). Our goal was to increase the pH of all saliva samples well above the indicator flip-point of pH 6.8, while not making the samples so basic that they couldn’t reach this pH upon successful target amplification. We found that human saliva containing 14.5 mM NaOH is optimal to inhibit false positives caused by saliva acidity (N=96; **Figure 1A**, right) without impeding the intended color change during amplification (**Figure 1B**). In addition, we designed our saliva stabilization solution to also include a chelating agent (1 mM EDTA final concentration) and Proteinase K to inhibit RNases, both of which help preserve virion RNA and therefore to increase sensitivity (note that Proteinase K will inhibit the RT-LAMP reaction if it does not go through a heat inactivation step prior to that reaction). Finally, the saliva stabilization solution contains TCEP, which aids in RNA stabilization by breaking disulfide bonds present in RNases and helping to reduce saliva viscosity. Our optimized saliva stabilization solution (2X solution: 5 mM TCEP, 2 mM EDTA, 29 mM NaOH, 100 μg/mL Proteinase K, diluted in DEPC-treated water) is key to this test. For additional advice on controlling the acidity of reactions see **Supplemental Text S1**.

### Optimized RT-LAMP primer sets for detecting SARS-CoV-2 in human saliva

A critical parameter in RT-LAMP is primer design because RT-LAMP requires 4-6 primers all working together (Notomi et al., 2000). We found that the “AS1E” set, developed by Rabe et. al. and targeting the *ORF1ab* region of the SARS-CoV-2 genome, performs very well (Rabe and Cepko, 2020). However, in order to target two distinct regions from the SARS-CoV-2 genome, we designed and tested a large number of additional primer sets. Two of our custom sets, “ORF1e” targeting the virus *ORF1ab* gene, and “CU-N2” targeting the virus *N* gene, exhibited similar sensitivity and amplification efficiency as the AS1E set, as determined using real-time fluorescence monitoring of RT-LAMP products (**Figure 2A, primer targeting sequences are highlighted in Supplemental S7**). We next confirmed that these primer sets were both compatible with saliva preserved in our saliva stabilization solution and with colorimetric RT-LAMP (**Figure 2B**).

**Figure 2:**
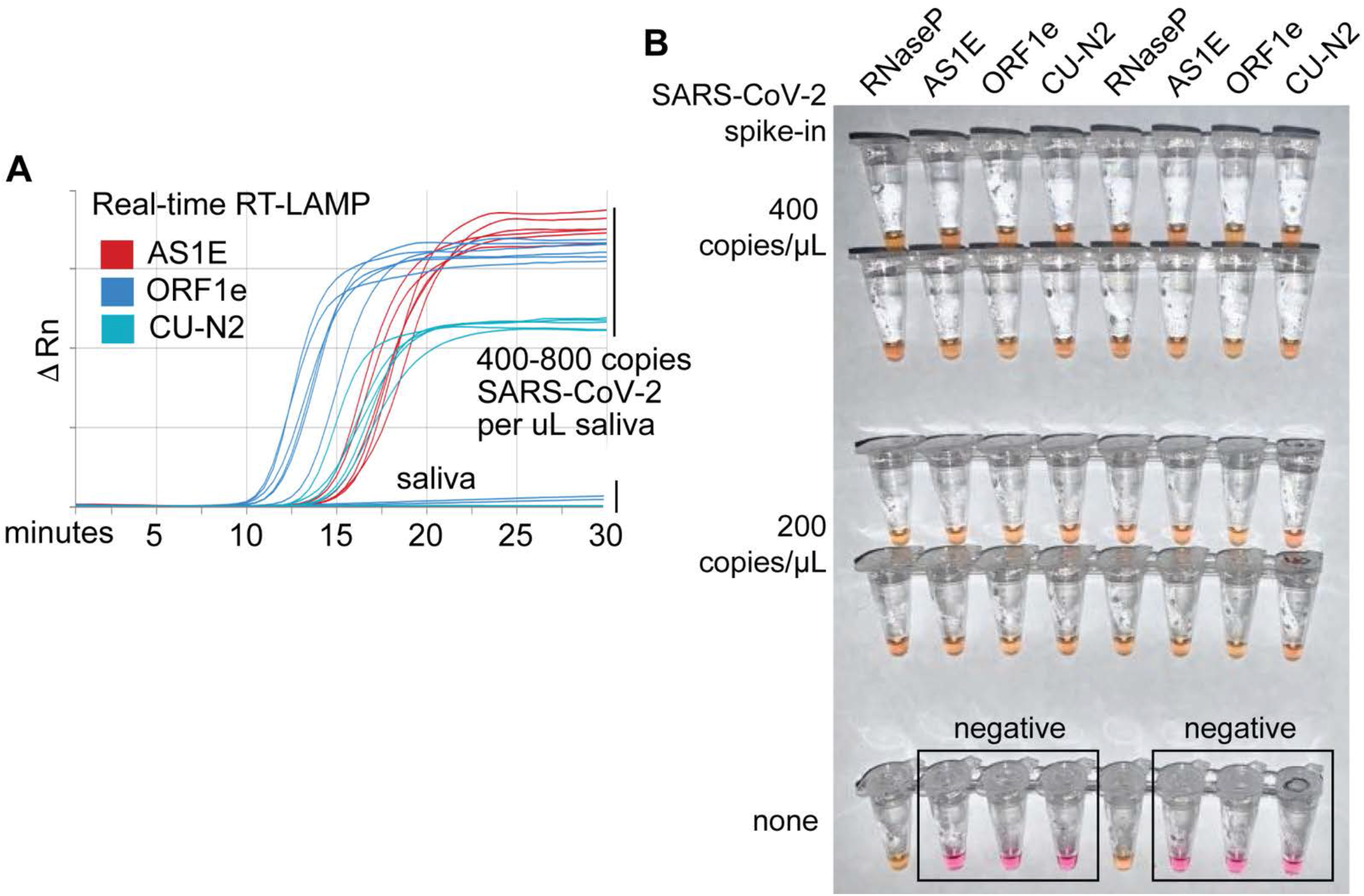
Optimized RT-LAMP primer sets for detecting SARS-CoV-2 in human saliva. **A)** Three RT-LAMP primer sets targeting the SARS-CoV-2 genome (AS1E (Rabe and Cepko, 2020), ORF1e, and CU-N2) were tested with real-time RT-LAMP. Saliva was mixed 1:1 with 2X saliva stabilization solution, heated at 95°C for 10 minutes, and then spiked with *in vitro* transcribed SARS-CoV-2 RNA at the indicated concentrations. 4 μL of this was added to a master mix containing primers and NEB’s WarmStart LAMP 2x Master Mix in a final reaction volume of 20 μL. Reactions were incubated at 65°C and a fluorescence reading was taken every 30 seconds. EvaGreen was used to monitor amplification products in real-time (X-axis) using a QuantStudio3 quantitative PCR machine. There are 9 lines for each of the three primer sets because three concentrations of spiked in SARS-CoV-2 RNA were each tested in triplicate (0, 400, 800 copies / μL saliva). The saliva samples without SARS-CoV-2 RNA spike in are shown as flat lines. When concentrations are given herein, denominator refers to the raw, pre-diluted saliva sample. The normalized change in fluorescence signal (ΔRn) is shown on the Y-axis. **B)** Saliva mixed 1:1 with 2X saliva stabilization solution was heated (95°C for 10 minutes) and then spiked with SARS-CoV-2 RNA at the indicated concentrations. Replicates were tested by RT-LAMP with the control RNaseP primer set and three distinct SARS-CoV-2 primer sets (AS1E, ORF1e, and CU-N2). All samples scored positive except those boxed, which are saliva samples that contain no SARS-CoV-2 RNA, as expected.

### Addressing biosafety concerns through heat inactivation

Next, we addressed the biosafety concerns of handling potentially infectious saliva samples. Recent studies suggest that incubation for 3 minutes at 95°C is sufficient to inactivate SARS-CoV-2 virions (Batéjat et al., 2021). However, when heating saliva samples for downstream analysis of RNA, one must balance heating long enough to liberate the target RNA from virions with not heating for so long that the target RNA will be degraded. Heating at 95 °C does degrade SARS-2-CoV RNA that is spiked directly into saliva samples but does not degrade viral RNA when it is spiked into samples within SARS-CoV-2 virions (**Supplemental Figure S1**). A 10-minute incubation of saliva samples at 95°C was found to be optimal (**Supplemental Figure S1**). We designed our test procedure such that testing personnel avoid handling open tubes until after this step to increase biosafety (**Supplemental Text S2**).

### Assessment of sample stability during storage

Stability of saliva samples from the time of collection to the time of processing and analysis is important if testing cannot be performed immediately, or if the tests are being conducted in batches. Saliva samples containing purified virions and diluted with 2X saliva stabilization solution were stored at 4°C for 24, 48, 72, or 96 hours before being inactivated at 95^°^C and analyzed using colorimetric RT-LAMP (**Supplemental Figure S2**). We tested saliva collection and storage over a range of SARS-CoV-2 virion spike-in concentrations. We observed no significant changes in sample stability and the test detection limit over this time course, suggesting that saliva samples stored in saliva stabilization solution at 4°C are stable for at least four days.

### Determining the limit of detection

We next sought to carefully evaluate the limit of detection for this test. The lowest concentration at which positive samples were reliably identified was 200 virions/μL in saliva (red box, summary table in **Figure 3A**). We next tested 20 replicates at this concentration (200 virions/μL) using all four primer sets (**Figure 3B**). The ORF1e primer set was not consistent in its performance at 200 virions/μL. Therefore, we decided to eliminate the ORF1e primer set from our testing panel and define a final colorimetric RT-LAMP test that includes primer sets RNaseP, AS1E, and CU-N2. Note that the limit of detection refers to the virus concentration that can be identified > 95% of the time, and the assay does often detect the virus at even lower concentrations.

**Figure 3:**
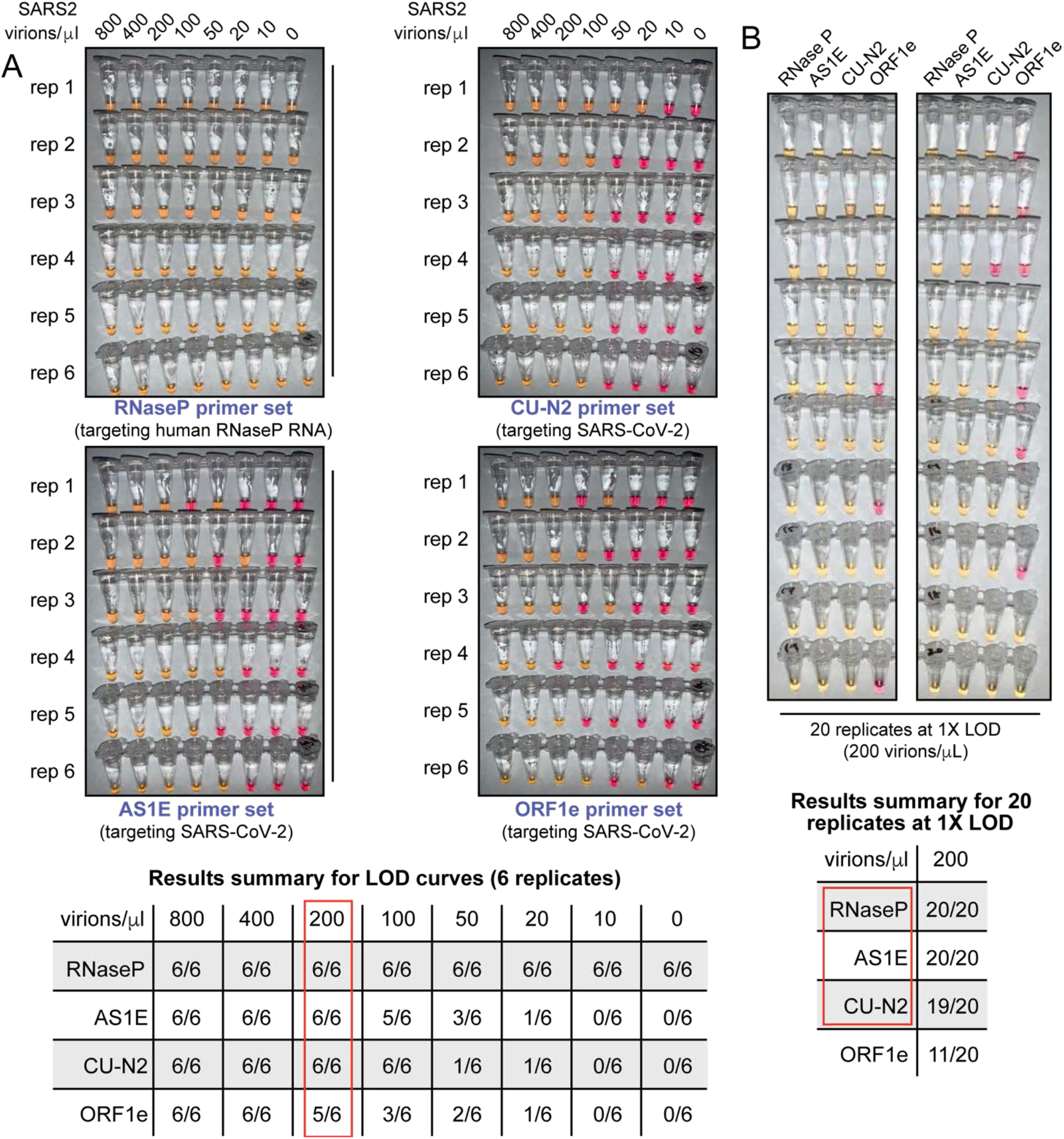
SARS-CoV-2 virion limit of detection using RT-LAMP and saliva samples. **A)** Saliva samples were spiked with the indicated concentrations of heat-inactivated SARS-CoV-2 virions (top) before being diluted 1:1 with 2X saliva stabilization solution. Samples were then heated at 95°C for 10 minutes and subjected to RT-LAMP at 65°C for 30 minutes in 6 replicates. Each panel represents a unique primer set (listed at the bottom of each panel). The table at the bottom shows a summary of positive reactions observed (yellow). Red box indicates the determined RT-LAMP limit of detection (LOD). **B)** Saliva samples were spiked with heat-inactivated SARS-CoV-2 virions at a concentration of 200 virions/μL (the limit of detection of our assay) before being diluted 1:1 with 2X saliva stabilization solution. Samples were then heated at 95°C for 10 minutes and 20 replicates of RT-LAMP with the indicated primer sets were incubated at 65°C for 30 minutes. The table at the bottom shows a summary of positive reactions (yellow). Red box indicates our selection of primer sets to advance to subsequent analysis.

We considered that contaminants in saliva and/or components of the saliva stabilization solution might be suppressing the overall RT-LAMP reaction efficiency by acting in inhibitory ways. On the contrary, we found that when synthetic SARS-CoV-2 RNA is directly added to the RT-LAMP reaction mix (in the absence of saliva and the stabilization solution), we were unable to achieve a better detection limit lower than 200 genome copies/μL (**Figure S3A**). This suggests the observed detection limit represents the upper performance limit of colorimetric RT-LAMP, and the saliva and stabilization solution have little to no negative impact to the test performance. In fact, multiple observations suggest that RNA degradation is observed in the absence of stabilization solution, resulting in less consistent testing results (**Figure 1A, Figure S2, Figure S3B**).

We next performed a blinded study. Heat-inactivated virions were spiked into human saliva at various concentrations at or above the limit of detection (200 virions/μL), and these as well as uninfected saliva samples were blinded and passed to a second member of our personnel. After running the RT-LAMP test on 60 such samples, only one positive sample scored as inconclusive. In that sample the SARS-CoV-2 primer set CU-N2 failed, while the other primer set detecting SARS-CoV-2 correctly identified the sample (**Supplemental Figure S4**). All negative samples were scored correctly (100% specificity, binomial 95% confidence interval [88%,100%]). Conservatively counting the inconclusive test as a false negative lead to a sensitivity estimate of 97% (binomial 95% confidence interval [93%,100%]). See **Figure 4C** for a breakdown by primer set.

**Figure 4.**
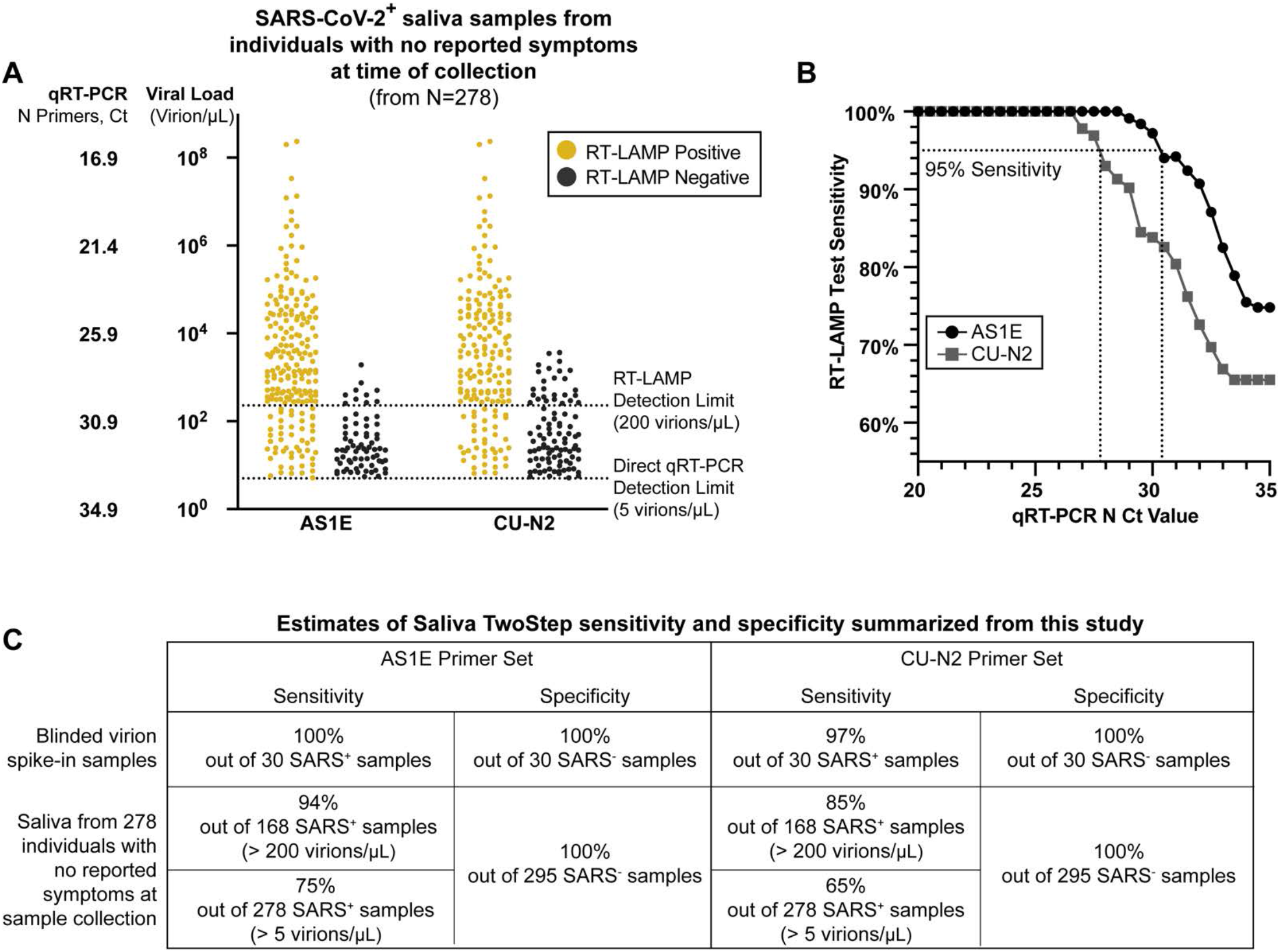
Evaluation of RT-LAMP on SARS-CoV-2-positive saliva samples from individuals with no reported symptoms at the time of sample collection. **A)** We re-analyzed university saliva samples that had been previously analyzed for SARS-CoV-2 using quantitative RT-PCR with a primer set against the N gene of SARS-CoV-2 (see methods). The remaining saliva was mixed 1:1 with 2X saliva stabilization solution (without Proteinase K) and re-tested using RT-LAMP. The results of RT-LAMP are compared to relative saliva viral load determined by quantitative RT-PCR. The figure shows the distribution of the viral load of all 278 positive saliva samples separated by the corresponding RT-LAMP reaction results with either the AS1E or CU-N2 primer-set. **B)** Saliva TwoStep RT-LAMP test sensitivity as a function of the cycle threshold (Ct) from the quantitative RT-PCR results of the corresponding SARS-CoV-2 positive saliva samples. **C)** A summary of the sensitivity and specificity of the Saliva TwoStep test from the blinded sample evaluation described above and shown in **Supplemental Figure S4** (top), and from both the data in panel A (bottom).

### Evaluation on human samples

SARS-CoV-2 screening was initiated on the University of Colorado Boulder campus starting in the summer/fall of 2020. Saliva samples were taken weekly from residents of dormitories and at several testing sites throughout the campus. Participants were asked to refrain from eating or drinking 30 minutes prior to sample collection, and to not participate if they were experiencing any symptoms consistent with COVID-19. These individuals were either pre-symptomatic at the time of saliva collection, or they never developed symptoms throughout the course of infection (we don’t have the necessary follow-up data to delineate these two outcomes). All saliva samples were first analyzed by a quantitative RT-PCR method performed directly on saliva mixed 1:1 with 2X TBE buffer containing 1% Tween-20 (Ranoa et al., 2020). An optimized multiplex quantitative RT-PCR reaction was used targeting the E and N gene regions of the SARS-2-CoV genome (see methods). From these, 295 negative samples and 278 positive samples were next re-evaluated with RT-LAMP. Each SARS-CoV-2-positive saliva sample has a Ct value associated with it from the quantitative RT-PCR test conducted by the campus testing team. Because positive results in our university screening regimen result in university affiliates being directed to their healthcare provider for confirmatory testing, with a few exceptions every positive sample is from a unique individual.

Saliva samples had already been heat inactivated for 30 minutes at 95°C as the initial step of the quantitative RT-PCR protocol. Since the heating component of our Saliva Preparation step had already been performed, an aliquot of the heated saliva sample was transferred into our 2X saliva stabilization solution (without Proteinase K) and then put through the RT-LAMP reaction as described above. For each of the 573 samples, three RT-LAMP reactions were performed with different primer sets: RNaseP (positive control), AS1E, and CU-N2 primer sets (the latter two sets detecting SARS-CoV-2). During this part of the study, we noticed that decreasing the input sample amount (saliva + saliva stabilization solution) from 4 μL to 2 μL in a total reaction volume of 20 μL further increases tolerance of the RT-LAMP reaction color to acidic saliva samples because less saliva is added. We thus reduced the input sample amount to 2 μL when evaluating these human samples. For all 573 samples, RT-LAMP with primers to human RNA positive control (RNaseP) correctly turned positive (yellow).

### Specificity

295 saliva samples that tested negative for SARS-CoV-2 by quantitative RT-PCR were used for evaluation. We re-tested all of those samples with RT-LAMP to evaluate our false-positive rate. Remarkably, for all 295 SARS-CoV-2-negative samples, AS1E and CU-N2 primers sets both universally returned a result of negative, consistent with the quantitative RT-PCR results. Therefore, there was zero false positive, and the test has a specificity of 100% in this extensive sample set. This shows the strength of our saliva stabilization solution, which mitigates the problem of false-positives in RT-LAMP due to some human saliva samples being naturally acidic.

### Sensitivity

We next analyzed 278 SARS-CoV-2-positive saliva samples with viral loads determined based on direct quantitative RT-PCR Ct values using a primer set directed against the nucleocapsid (N) gene of SARS-CoV-2 (see methods). All Ct values reported in this study are from this primer set. We determined the relative viral load of each positive saliva sample based the quantitative RT-PCR standard curve generated by our university testing lab (**Supplemental Figure S5**). Among all positive samples, 208 (74.8%; AS1E primers) and 182 (65.5%; CU-N2 primers) returned positive RT-LAMP test results (**Figure 4A**). Although both primers sets were still able to detect SARS-CoV-2 RNA below the experimentally determined detection limit (200 virions/μL), we observed a decline in the test sensitivity below such limit (**Figure 4B**). Of the 168 positive samples that contain greater viral load than RT-LAMP limit of detection, 158 (94%; AS1E primers) or 142 (85%; CU-N2 primers) returned positive RT-LAMP test results (**Figure 4A**). In **Figure 4C**, we summarize the performance of each primer set in both this test of human saliva samples and in the spiked in virion experiments described above (**Supplemental Figure S4**). The observed limit of detection of the AS1E primer set was determined from this data to be 266 virions/microliter. The strong congruence with our prior estimate of 200 virions/microliter demonstrates that heating for 30 minutes prior to adding stabilization solution and using 2 µL of saliva plus stabilization solution, instead of 4 µL, both have very little effect. Because the AS1E primer set performs best throughout our study, we include that as the main primer set in our final test configuration, which we refer to as the Saliva TwoStep test. However, the CU-N2 primer set still performs well and can be used when it is desirable to detect a second region of the SARS-CoV-2 genome.

### Test sensitivity as a function of viral load in the sample

For both primer sets, we calculated the sensitivity (positive agreement with quantitative RT-PCR) and specificity (negative agreement with quantitative RT-PCR) of the RT-LAMP test at various levels of viral load cutoffs (**Figure 4B, Table 1**). The differences in the observed limit of detection between the two SARS-CoV-2 primer sets could reflect the differences in the primer efficiencies, as well as the dynamics in relative viral transcript abundance(Kim et al., 2020).

**Table 1.** Summary of RT-LAMP evaluation in human samples.

|  |  |  | RT-LAMP |  |  |  |  |
| --- | --- | --- | --- | --- | --- | --- | --- |
|  |  |  | No. of Samples | AS1E |  | CU-N2 |  |
|  |  |  |  | No. of Positives | Agreement | No. of Positives | Agreement |
| Quantitative RT-PCR (SARS-CoV-2 N) | Negative |  | 295 | 0 | 295/295 (100%) | 0 | 295/295 (100%) |
| | Positive<br>(Levels of Viral Load: Virions/μL) | 4000 | 82 | 82 | 82/82 (100%) | 82 | 82/82 (100%) |
|  |  | 2000 | 97 | 97 | 97/97 (100%) | 94 | 94/97 (96.9%) |
|  |  | 1000 | 118 | 117 | 117/118 (99.2%) | 110 | 110/118 (93.2%) |
|  |  | 800 | 123 | 122 | 122/123 (99.2%) | 112 | 112/123 (91.1%) |
|  |  | 400 | 143 | 139 | 139/143 (97.2%) | 129 | 145/173 (90.2%) |
|  |  | 200 | 168 | 158 | 158/168 (94.0%) | 142 | 142/168 (84.5%) |

### Assessment of Saliva TwoStep against an EUA approved nasal swab test

Of the 278 SARS-CoV-2-positive saliva samples analyzed above, 54 also had matched nasal samples collected no more than two days later. In some cases, individuals may have developed symptoms by the time follow-up nasal swabs were taken, so we can made no claims about symptomatic status at the time of nasal swab. We next compared the results of the Saliva TwoStep test with the result obtained by the Quidel Lyra direct nasal swab RT-PCR test. Compared to the quantitative RT-PCR on saliva results, the RT-LAMP produced three false negative in this sample set, whereas the Lyra nasal swab test produced eight (**Figure 5A**). However, this is still remarkably consistent given that this comparison involves three degrees of freedom: biosample (saliva versus nasal swab), test modality (RT-PCR versus RT-LAMP), and days between saliva and nasal samples collection (up to two days apart). A summary of how these first two degrees of freedom affect test congruency are shown in **Figure 5B**.

**Figure 5.**
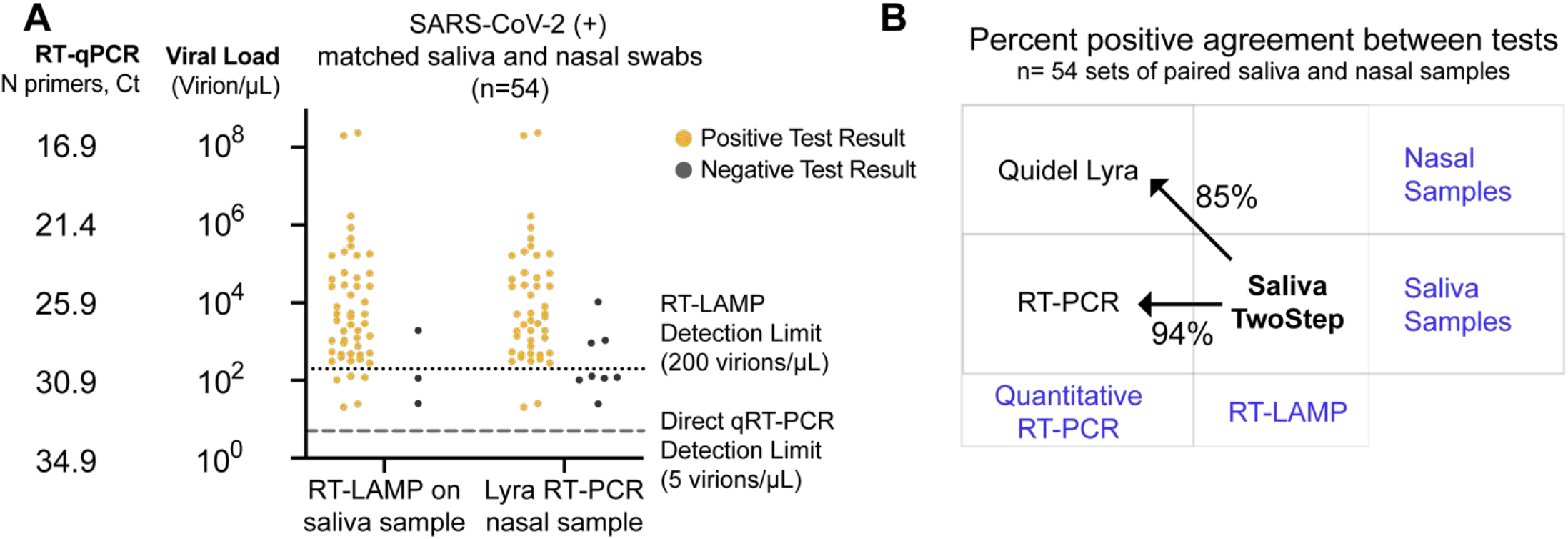
Assessment of Saliva TwoStep against an EUA approved nasal swab test. **(A)** Matched nasal swabs and saliva from 54 individuals were analyzed (all of whom were SARS-CoV-2 positive at the time that these samples were collected, as verified by saliva direct quantitative RT-PCR test). Nasal swab samples from the same individuals were collected within 2 days of positive saliva test, and tested using the EUA Quidel Direct Lyra RT-PCR test. The saliva samples from those same individuals were tested with the Saliva TwoStep test. Data points represent individuals (n = 54), and the corresponding test result is color-coded: positive, yellow; negative, grey. **(B)** Positive test agreement between Saliva TwoStep and the two comparator tests. The nature of the sample used by each test (nasal swab or saliva), and the test chemistry (quantitative RT-PCR or RT-LAMP) are delineated.

### Final test conditions

From the experiments described above we selected the final optimized conditions for our Saliva TwoStep test. The two steps have an end-to-end processing and analysis time of approximately 45 minutes (**Figure 6**). For additional application details regarding the testing station setup, sample collection, and overall workflow of employing this test for community screening, please refer to the Supplemental Text S1 and Supplemental Text S2.

**Figure 6.**
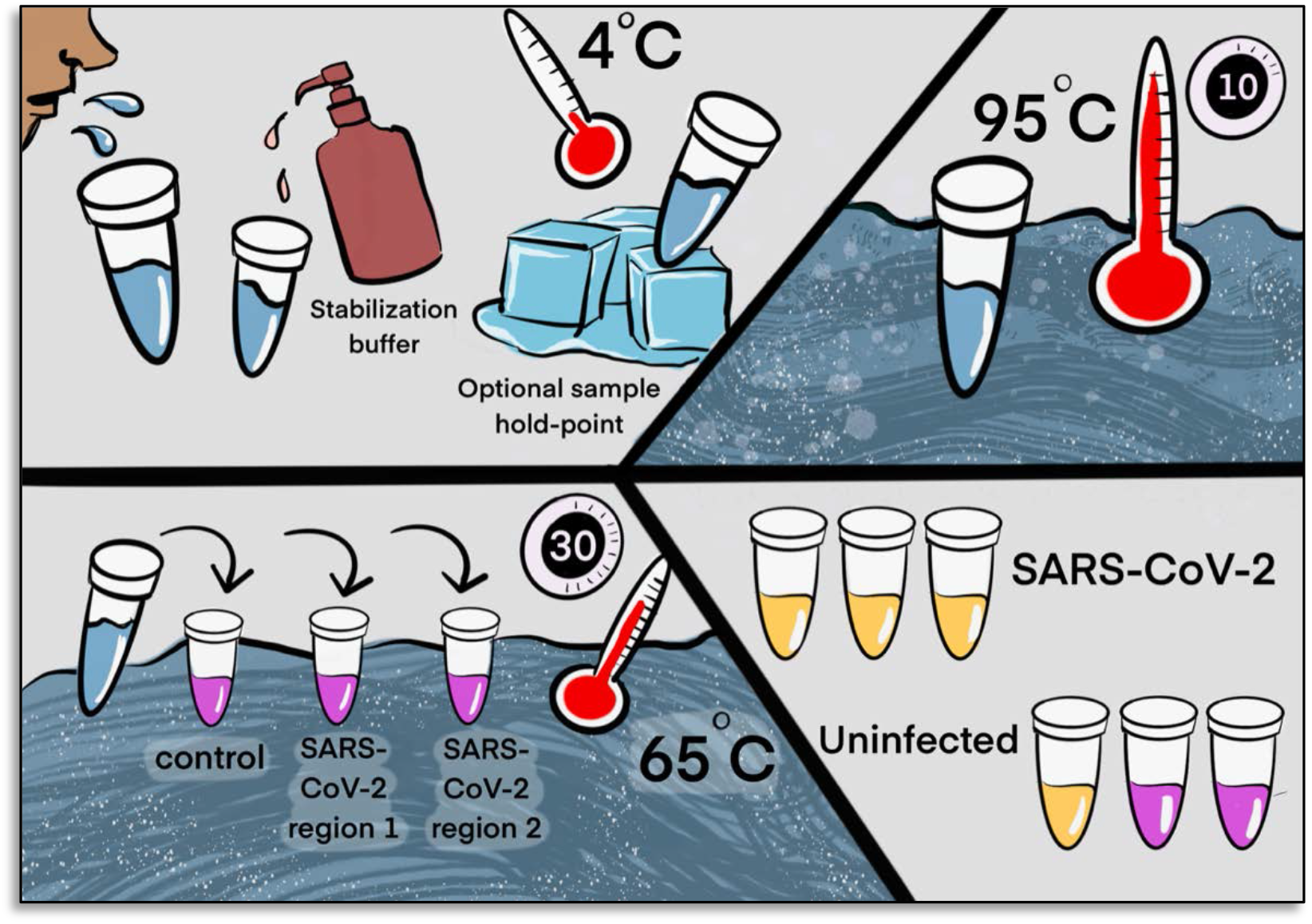
Two step detection of SARS-CoV-2 in saliva. Upper half) Step 1: Prepare Saliva. Person provides 1 mL of saliva, and 1 mL of 2X saliva stabilization solution is then added to it. (This sample can be processed immediately or stored in the refrigerator at 4^°^C for at least 4 days.) The mixture is heated at 95^°^C for 10 minutes. This step serves to neutralize the pH of saliva, liberate viral RNA from virions in the saliva, and inactivate virions for safe handling (although appropriate safety precautions should always be taken). We have determined that performing a heating step at 95^°^C for 30 minutes in a water bath before addition of the saliva stabilization solution also works equally well. However, in this case Proteinase K must be left out of that solution. **Lower half) Step 2: Detect Virus**. 2 μL of stabilized saliva from step 1 is pipetted into each of three test tubes pre-filled with the RT-LAMP master mix and primers. The only thing different between the three tubes is the primer set included, with each set targeting either the human positive control RNA or a region of SARS-CoV-2 RNA, as indicated. After incubation, the reaction will turn from pink to yellow if the target RNA is present in saliva. An example of a positive and a negative test are shown. Graphic by Annika Rollock.

#### Step 1. Prepare Saliva

Collect saliva, combine 1:1 with 2X saliva stabilization solution and incubate at 95°C for 10 minutes. *Note:* We have determined that performing a heating step at 95^°^C for 30 minutes in a water bath, before addition of the saliva stabilization solution, also works reasonably well. However, in this case Proteinase K must be omitted.

#### Step 2. Detect Virus

Incubate at 65°C for 30 minutes: 2 μL diluted saliva from step 1, 10 μL 2X NEB Colorimetric RT-LAMP enzyme mix, 6 μL of nuclease-free water and 2 μL 10x primer mix for a final reaction volume of 20 μL.

#### Step 3. Reaction Inactivation *(optional)*

Stop reaction at 80°C for 2 minutes. This stabilizes color so that results can be analyzed at a later time. The multiple heating steps here may be programmed into a thermal cycler for maximum convenience, but this is not necessary.

## Discussion

There are several advantages to the SARS-CoV-2 Saliva TwoStep RT-LAMP screening approach described herein: **1)** The use of saliva eliminates invasive nasal swab-based sampling, which requires special supplies and causes discomfort. **2)** We optimized saliva stabilization solution that allows for the neutralization of a broad range of naturally acidic saliva samples while maintaining compatibility with a colorimetric RT-LAMP assay. The solution also helps preserve saliva samples for at least four days before processing and lowers saliva viscosity. **3)** We determined the optimal sample heating condition that liberates the host and viral RNA with minimal degradation. The simple heating step increases biosafety and avoids formal RNA extraction procedures. **4)** For RT-LAMP, we incorporated additional primers based on up-do-date SARS-CoV-2 genome databases and identified primers allowing efficient target amplification. These primers are expected to work on most of all viral variants currently circulating (**Supplemental Figure S7)**. Overall, with the simplified two steps of saliva preparation and virus detection, the test has a rapid sample-to-result turnaround time of 45 minutes.

Through the optimization process, we identified other potential sources of false positive results and provided a detailed summary for troubleshooting (**Methods and Supplemental Text S1**). In addition, from our experience of the actual deployment of this screening test, we summarized the standard operational procedures for saliva sample collection, including the design of a stabilization solution dispensing apparatus to preserve samples while avoiding environmental contamination and protecting workers (**Supplemental Figure S6, Supplemental Text S2**). By strictly following these application notes, we completely avoided false positive results during the evaluation of a large cohort of human saliva samples, achieving 100% specificity. We also evaluated the RT-LAMP test performance based on the experimentally determined limit of detection. Using SARS-CoV-2 positive human saliva samples, we confirmed that the RT-LAMP test can consistently identify infected individuals with 94% sensitivity.

During the test development and optimization, we have also explored additional methods that may help enhance the RT-LAMP test performance and consistency. Previous work suggests that the addition of 40 mM of guanidine chloride in the RT-LAMP reaction mix could increase RT-LAMP amplification efficiency (Zhang et al., 2020). However, we did not observe similar enhancement when included in our experiments. To further prevent carry over contamination (Davidi et al., 2020), the usage of dUTP and uracil-DNA-glycosylase-containing RT-LAMP reaction mix can be considered. Through data not shown, we determined that the addition of this alternative master mix does not affect the reported test limit of detection.

Saliva TwoStep requires less sample processing, reaction incubation time, and laboratory overhead as compared to quantitative RT-PCR methods. The result is the ability to run significantly more tests with a given amount of resources. Based on these observations, we conclude that the Saliva TwoStep test described herein can be used as a SARS-CoV-2 screening tool to reliably identify highly infective individuals with minimal laboratory setup, potentially serving as a tool for effective SARS-CoV-2 surveillance at the community level. This RT-LAMP testing offers many solutions to a nation-wide shortage of COVID-19 testing. With minimal set-up this test could be performed in diverse settings such as factories, office buildings, or schools.

### Ethics statements

This study was approved by the University of Colorado Boulder Institutional Review Board. Saliva samples for assay development were collected under protocol 20-0068. Testing on human subjects is aggregated data resulting from University of Colorado Boulder operational COVID-19 surveillance testing.

## Data Availability

All data is included in the manuscript.

## Acknowledgements

We thank Dan Larremore for valuable advice, Annika Rollock for help with graphics, and the University of Colorado Boulder for their ambitious and aggressive approach to keeping our campus safe. In particular, we thank Kristen Bjorkman for playing a key part in making it all happen. We dedicate this study to the memory of our wonderful colleague, Denise Muhlrad.

## Material and Methods

### RT-LAMP primer design and preparation

Regions of the SARS-CoV-2 genome that are conserved among strains were identified using genome diversity data from NextStrain (nextstrain.org/ncov/global). Next, nucleotide-BLAST (blast.ncbi.nlm.nih.gov) was used to filter out genome sequences that share high sequence homology with other seasonal coronavirus genomes. Finally, PrimerExplorer V5 (<u>primerexplorer.jp/e/</u>) was used to design RT-LAMP primers targeting the specific regions of SARS-CoV-2 genomes. The F3, B3, FIP, BIP, Loop F and Loop B primers were selected for optimal melting temperature and complementarity using A plasmid editor (ApE). All primers were ordered from IDT in desalted form. In all cases, a 10X concentration of primer sets was made containing 16 μM FIP and BIP primers, 4 μM LF and LB primers, and 2 μM F3 and B3 primers.

All primers should be ordered with HPLC purification, which ensures the yield and avoids cross contamination from other SARS-CoV-2-related synthesis projects being run on the same equipment at the oligo synthesis facilities (which can lead to false positives). This is particularly a problem during a pandemic where these facilities are handling many oligo synthesis orders focused on the same pathogen(Mögling et al., 2020). It is also recommended that you communicate with the primer synthesis company to inform them that primers are intended for use with a SARS-CoV-2 screening test. Several companies have dedicated facilities for minimizing cross-contamination of SARS-CoV-2 templates. In addition, primers should be diluted in nuclease-free water, instead of Tris-EDTA (TE) buffer, which will also inhibit pH change that takes place during RT-LAMP.

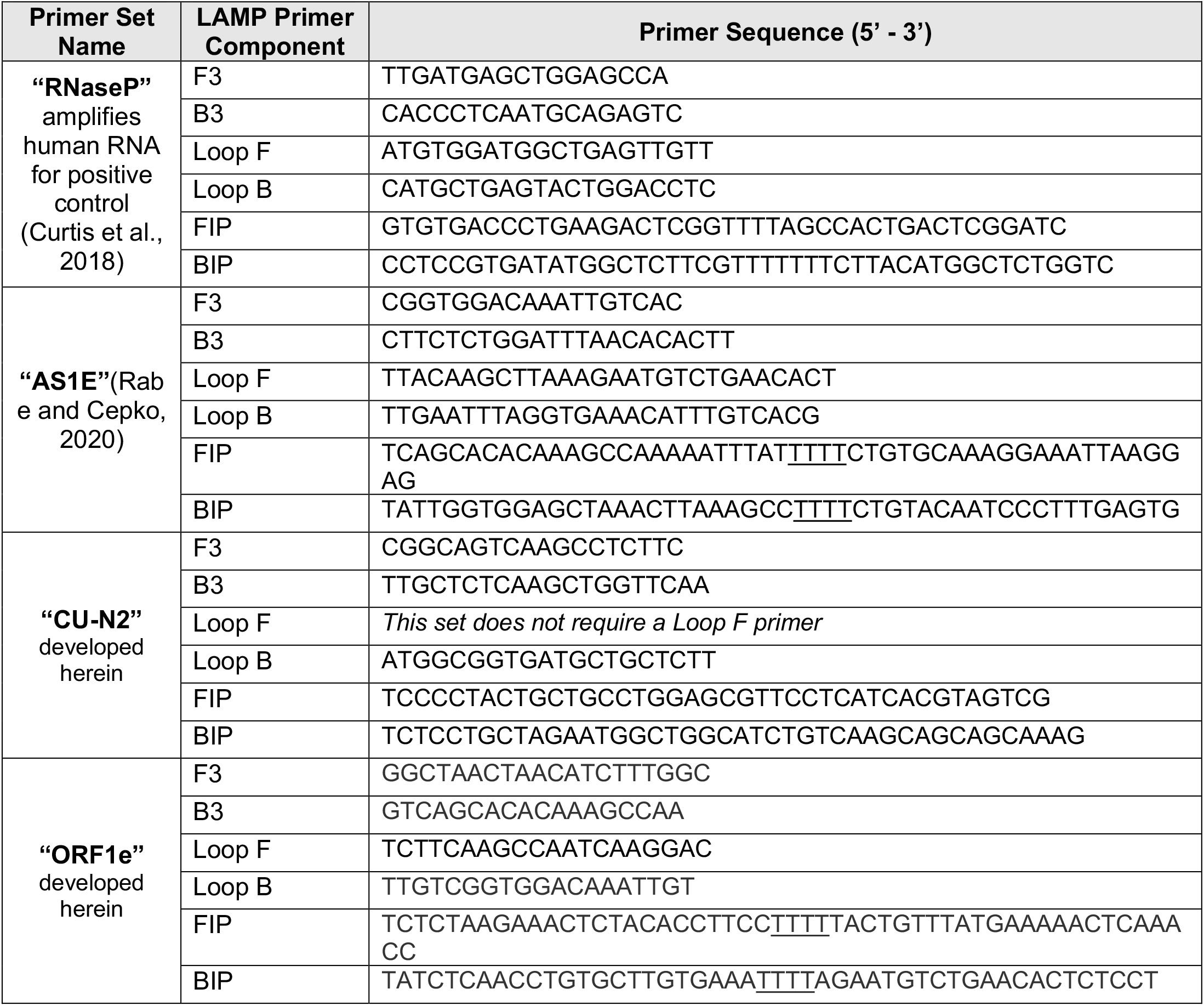

### SARS-CoV-2 RNA and virion standards

Synthetic SARS-CoV-2 RNA control (Twist Bioscience #102019) was obtained and its copy number of 1×10^6^ copies/μL was confirmed using quantitative RT-PCR in conjunction with a DNA plasmid control containing a region of the N gene from the SARS-CoV-2 genome (IDT #10006625). Heat-inactivated SARS-CoV-2 virion control (ATCC #VR-1986HK) was obtained and its concentration of 3.75×10^5^ virions/μL was confirmed using quantitative RT-PCR in conjunction with both the synthetic SARS-CoV-2 RNA control and a DNA plasmid control containing a region of the N gene from the SARS-CoV-2 genome. SARS-CoV-2 RNA was added to saliva samples after being mixed 1:1 with saliva stabilization solution and heated at 95°C for 10 minutes, unless stated otherwise, whereas heat-inactivated SARS-CoV-2 virions were added to saliva samples and mixed 1:1 with saliva stabilization solution before being heated. Concentrations reported throughout this study represent the final concentration of standards in saliva before it was mixed 1:1 with 2X saliva stabilization solution.

### Saliva preparation with heat and stabilization solution

When making the 2X saliva stabilization solution, we offer several key pointers: 1) Use TCEP-HCl (GoldBio #TCEP10). The -HCL form must be used to produce the correct final stock pH. 2) Use EDTA, 0.5 M, pH 8.0 (Sigma-Aldrich #324506). It is important to use a pH 8.0 stock solution, otherwise this also affect the pH of the final stabilization solution. 3) Use lyophilized Proteinase K (Roche # 3115879001). It is important to use the lyophilized form. Liquid forms will contain Tris, which inhibits the pH change during the RT-LAMP reaction. 4) 10 M NaOH was prepared by dissolving NaOH pellets (Sigma-Aldrich #221465) into nuclease-free water, before being added to the 2X solution to reach the correct concentration. The following is the exact recipe that we used to create a 100mL stock of 2X saliva stabilization solution:

| Components | Amount Mixed | Final Concentration in 2X stock solution |
| --- | --- | --- |
| TCEP-HCl (GoldBio #TCEP10) | 143.3 mg | 5 mM |
| 0.5 M EDTA, pH 8.0 (Sigma-Aldrich #324506) | 400 µL | 2 mM |
| 10 M NaOH (Sigma-Aldrich #221465) | 290 µL | 29 mM |
| Proteinase K (Roche #3115879001) | 10 mg | 100 µg/mL |
| Nuclease-free water | To 100 mL |  |

Saliva samples (1 mL) were collected in sterile, nuclease-free 5 mL conical screw-cap tubes (TLD Five-O # TLDC2540). 2X saliva stabilization solution described above was then added at a 1:1 ratio. Samples were shaken vigorously for 5 -10 seconds and incubated at 95°C for 10 minutes. Samples were then placed on ice before being used in downstream analyses (Detailed sample collection procedure is described in **Supplemental Text S2**).

### Real-time RT-LAMP

For each reaction, 10 μL WarmStart LAMP 2X Master Mix (NEB #E1700) was combined with 1 μL 20X EvaGreen Dye (Biotium #31000), 2 μL 10X primer mix, and 3 μL DEPC-treated water. The combined reaction mix was added to MicroAmp Optical 96-Well Reaction Plate (ThermoFisher #N8010560) and then 4 μL processed saliva sample was added. The reaction was mixed using a multi-channel pipette and incubated in Applied Biosystems QuantStudio3 Real-time PCR system. The reaction proceeded at 65°C for 30 minutes with fluorescent signal being captured every 30 seconds. The results were visualized and analyzed using ThermoFisher’s Design and Analysis software.

### Colorimetric RT-LAMP

WarmStart Colorimetric LAMP 2X Master Mix (NEB #M1800) was used in all colorimetric RT-LAMP reactions. Each reaction was carried out in a total of 20 μL (10 μL WarmStart Master Mix, 2 μL 10X primer mix, 4 μL processed saliva sample, and 4 μL DEPC-treated water). Reactions were set up in PCR strip tubes on ice. Saliva template was added last and tubes were inverted several times to mix samples and briefly spun down in a microfuge. Reactions were incubated in a thermal cycler at 65°C for 30 minutes and then deactivated at 80°C for 2 minutes. The incubation was carried out without the heated lid to simulate a less complex heating device. Images of reactions were taken using a smartphone. For the community deployment of this assay, 2 μL of processed saliva was used instead of 4 μL.

### Testing of University Samples

The University of Colorado Boulder SARS-CoV-2 screening test was loosely based on a published quantitative RT-PCR reaction performed directly on saliva (Ranoa et al., 2020), which has a limit of detection of 5 virions/μL. Some modifications were made, as described here. For sample collection, individuals were asked to collect no less than 0.5 mL of saliva in a 5 mL screw-top collection tube. Saliva samples were heated at 95^°^C for 30 minutes to inactivate the viral particles for safe handling, and then placed on ice or at 4^°^C. For quantitative RT-PCR analysis, the university testing team transferred 75 μL of saliva into a 96-well plate, where each well had been pre-loaded with 75 μL 2xTBE buffer supplemented with 1% Tween 20. (The remaining saliva in the 5 mL collection tube proceeded to RT-LAMP testing as described in the next paragraph). Next, 5 μL of this diluted sample was added to a separate 96-well plate containing 15 μL reaction mix composed of: TaqPath 1-step Multiplex Master Mix (Thermo Fisher A28523), nuclease-free water, and triplex primer mix consisting of primer and probe sets targeting SARS-CoV-2 E and N genes and human RNase P gene (sequence and concentration specified in the table below). The reactions were mixed, spun down, and loaded onto a Bio-Rad CFX96 or CFX384 qPCR machine. Quantitative RT-PCR was run using the standard mode, consisting of a hold stage (25^°^C for 2 minutes, 50^°^C for 15 minutes, and 95^°^ C for 2 minutes) followed by 44 cycles of a PCR stage (95^°^ C for 3 seconds, 55^°^C for 30 seconds, with a 1.6^°^C/sec ramp up and ramp down rate). Only Ct values from the N primer set are reported in the study herein, and used to calculate relative sample viral load based on the standard curve shown in **Supplemental Figure S5**.

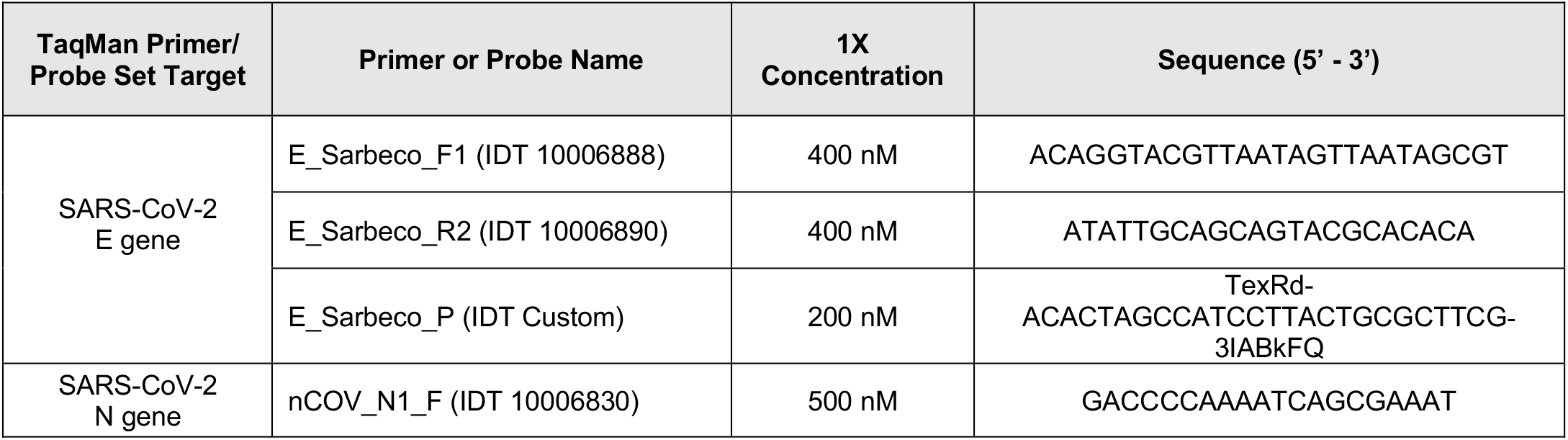
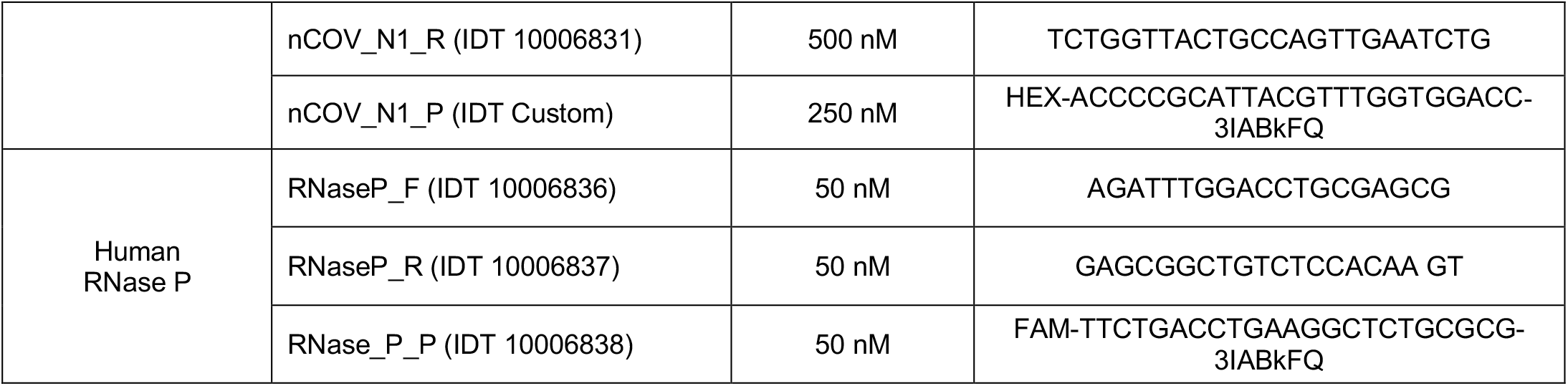

Leftover samples from this testing procedure were then tested with RT-LAMP. 50 μL of saliva samples was transferred and mixed into a 96-well plate, pre-loaded with 50 μL 2X saliva stabilization solution without proteinase K (5 mM TCEP-HCL, 2 mM EDTA, 29 mM NaOH, diluted in DEPC-treated water). 2 μL of diluted saliva samples were transferred into 8-strip PCR tubes containing RT-LAMP reaction mixture (enzymes and primers). For each sample, three RT-LAMP reactions were carried out to amplify human RNaseP as a control and AS1E and CU-N2 for SARS-CoV-2. The reactions were incubated at 65°C for 30 minutes followed by inactivation at 80°C for 2 minutes on a thermal cycler (Bio-RAD T100). A color change from pink to yellow was observed visually to interpret results.

**Figure S1:**
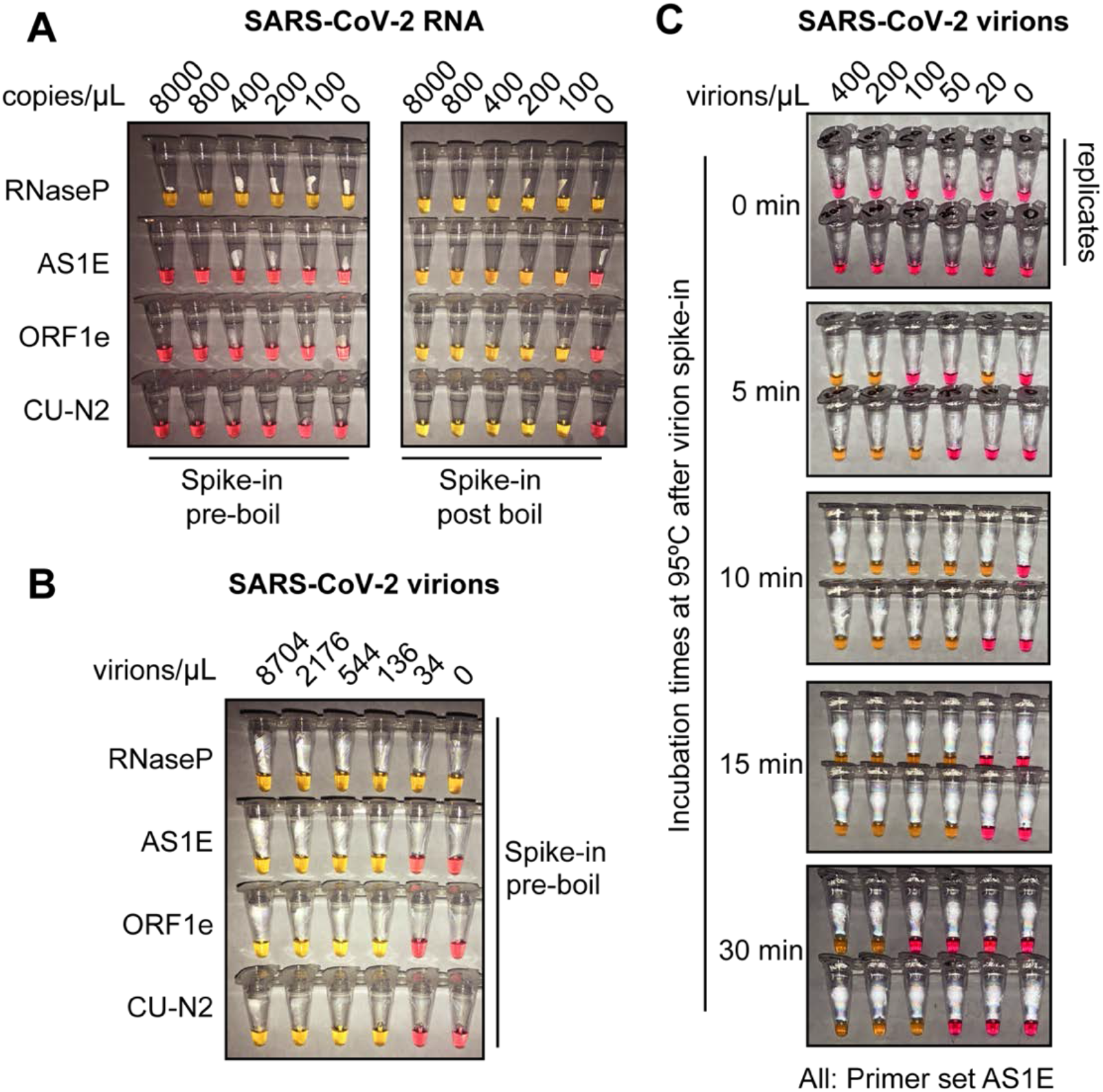
Optimized heat inactivation for safely detecting SARS-CoV-2 in human saliva. **A)** This experiment shows that heating at 95^°^C for 10 minutes degrades viral RNA when it is not in the form of virions. Saliva samples were diluted 1:1 with saliva stabilization solution. *In* vitro transcribed SASR-CoV-2 RNA was spiked into the diluted saliva to reach the indicated concentrations before (left) or after (right) the heating at 95°C for 10 minutes. To match other experiments, the indicated concentration represents the copies of SASR-CoV-2 RNA in the original undiluted saliva. The samples were then subjected to RT-LAMP at 65°C for 30 minutes. In this colorimetric version of RT-LAMP, reactions remain pink when no amplification occurred, and turned yellow if there was an amplification event. An RT-LAMP primer set targeting the human RNaseP transcript is included as a host RNA amplification control in addition to the three SARS-CoV-2 primer sets shown in panel A. **B)** This experiment shows that heating saliva at 95^°^C for 10 minutes does not degrades viral RNA when it is in the form of virions. Saliva samples were spiked with the indicated concentrations of heat-inactivated SARS-CoV-2 virions before being diluted 1:1 with saliva stabilization solution. Samples were then heated at 95°C for 10 minutes and subjected to RT-LAMP similarly to the experiment shown in panel A. **C)** Results illustrate the optimal incubation time at 95^°^C to liberate SARS-CoV-2 RNA from virions. Saliva samples were spiked with the indicated concentrations of heat-inactivated SARS-CoV-2 virions before being diluted 1:1 with saliva stabilization solution. Samples were then heated at 95°C for the indicated amount of time, and subjected to RT-LAMP similarly to the experiment shown in panel B. Without heating, no SARS-CoV-2 RNA can be detected with RT-LAMP, presumably because virions remain intact and the viral RNA is not accessible by the amplification enzymes. Amplification is somewhat inconsistent at 5 and 30 minutes possibly because at 5 minutes hardly any RNA has been liberated, and by 30 minutes it has been largely degraded. However, 10 or 15 minutes at 95^°^C appears to provide just the right balance between liberating and preserving RNA. All reactions contain the AS1E primer set. Duplicates are presented at each time point.

**Supplemental Figure S2:**
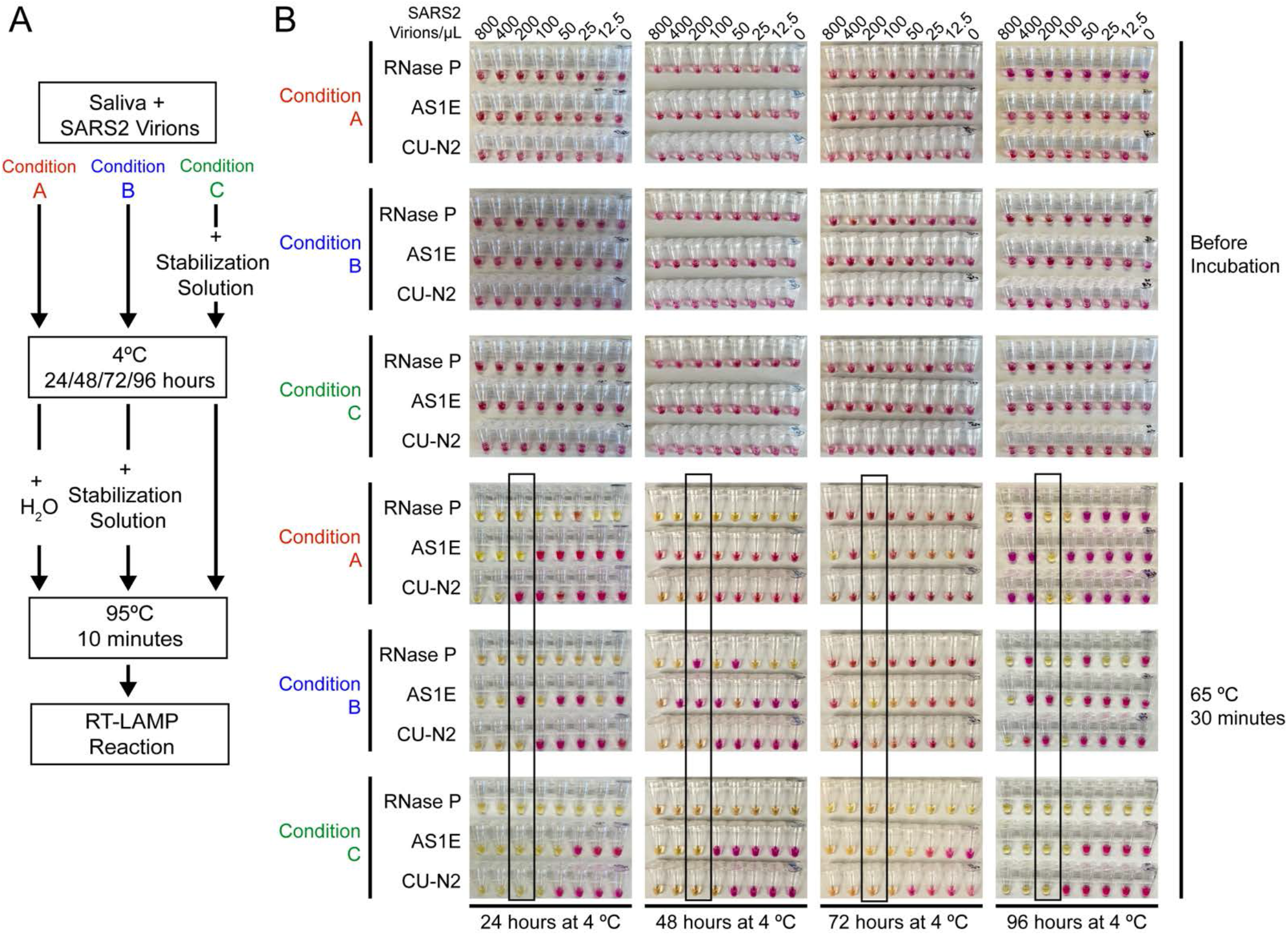
Saliva samples are stable at 4°C for at least 4 days before processing, if stored in saliva stabilization solution. **A)** Schematic of the experimental conditions. **B)** RT-LAMP reaction result before and after the isothermal amplification. Saliva samples were spiked with heat-inactivated SARS-CoV-2 virions at the indicated concentration and mixed 1:1 with saliva stabilization solution or nuclease-free water before/after storing at 4 °C for 24, 48, 72 and 96 hours. Samples were then heated at 95°C for 10 minutes and analyzed using RT-LAMP with the indicated primer sets. Condition C, which is the condition used in our test, performs robustly and is sensitive to the limit of detection even after 96 hours storage at 4^°^C. The stated limit of detection of 200 virions/μL is boxed.

**Supplemental Figure S3:**
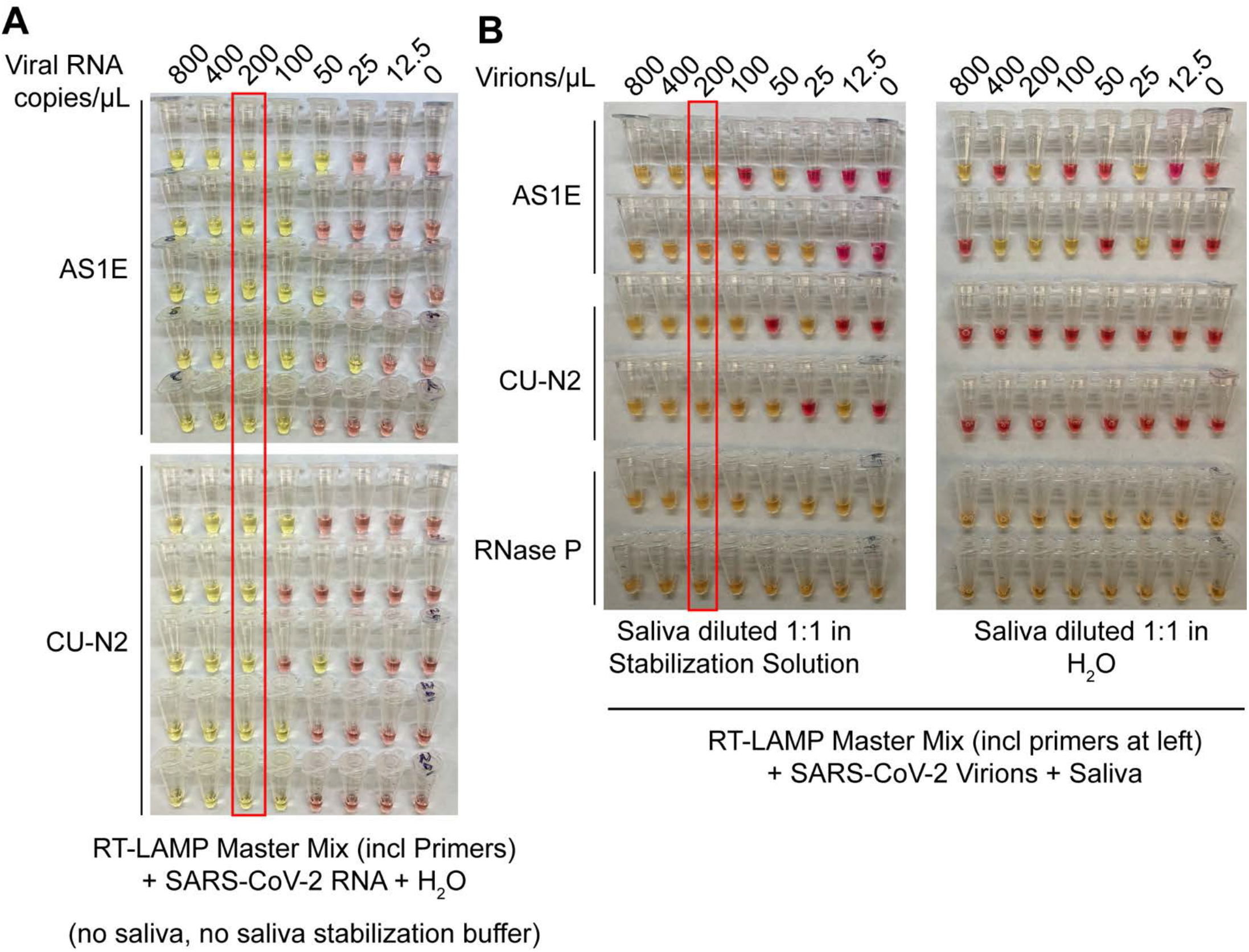
Saliva stabilization solution containing NaOH does not lower sensitivity of colorimetric RT-LAMP detection of SARS-COV-2. **A)** Here, the detection limit of Saliva TwoStep RT-LAMP assay, in the absence of any saliva or saliva stabilization solution, was assessed. This was explored in order to determine whether there might be components of saliva or saliva stabilization solution that inherently lower test sensitivity because they are inhibitory to the RT-LAMP reaction. Here, synthetic SARS-CoV-2 RNA was diluted in nuclease-free water. The diluted RNA was mixed with RT-LAMP reaction mix and incubated at 65°C for 30 minutes to allow isothermal amplification. Positive reactions turn yellow. Two different primer sets that amplify SARS-CoV-2 were employed, AS1E and CU-N2. The red box indicates the concentration at which positives were identified at least 95% of the time (here, 100% is achieved). That is defined at the limit of detection. Here, it is 200 copies/μL, just as when saliva and saliva stabilization solution is used (see panel B, and data figures in main paper). **B)** Evaluation of RT-LAMP detection limit in the presence of saliva, but in the presence or absence of saliva stabilization solution. Saliva spiked with heat inactivated SARS-CoV-2 virions at specified concentrations was mixed 1:1 with stabilization solution (left) or nuclease-free water (right) and heated at 95°C for 10 minutes (RNA liberation) before being incubated at 65°C for 30 minutes (isothermal amplification). On the left, the saliva stabilization solution achieves a limit of detection of 200 virions /μL. When virions are boiled without the saliva stabilization solution (right), very few reactions turn positive and the pattern is unpredictable, presumably because virions and viral RNA are destroyed.

**Supplemental Figure S4:**
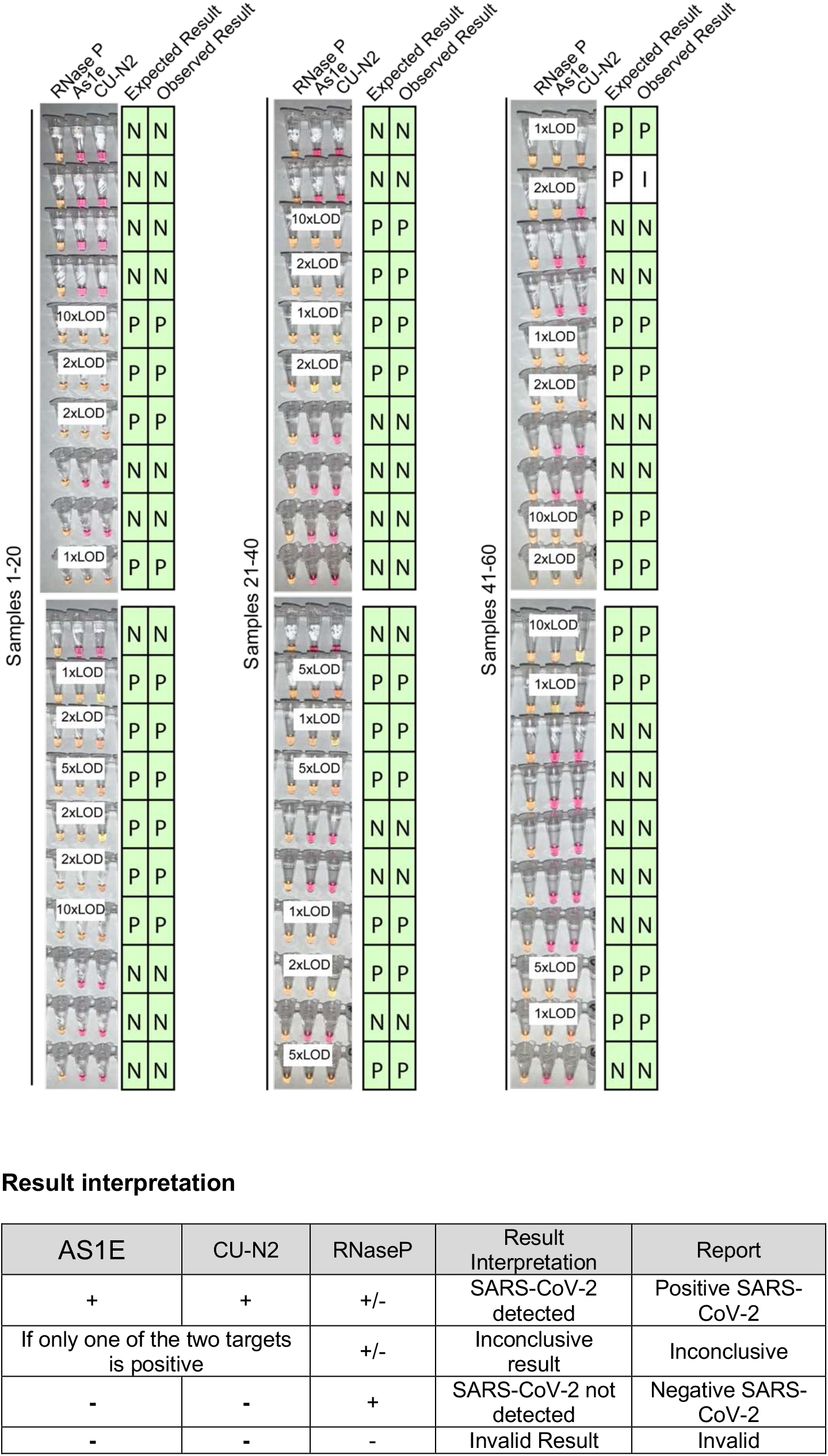
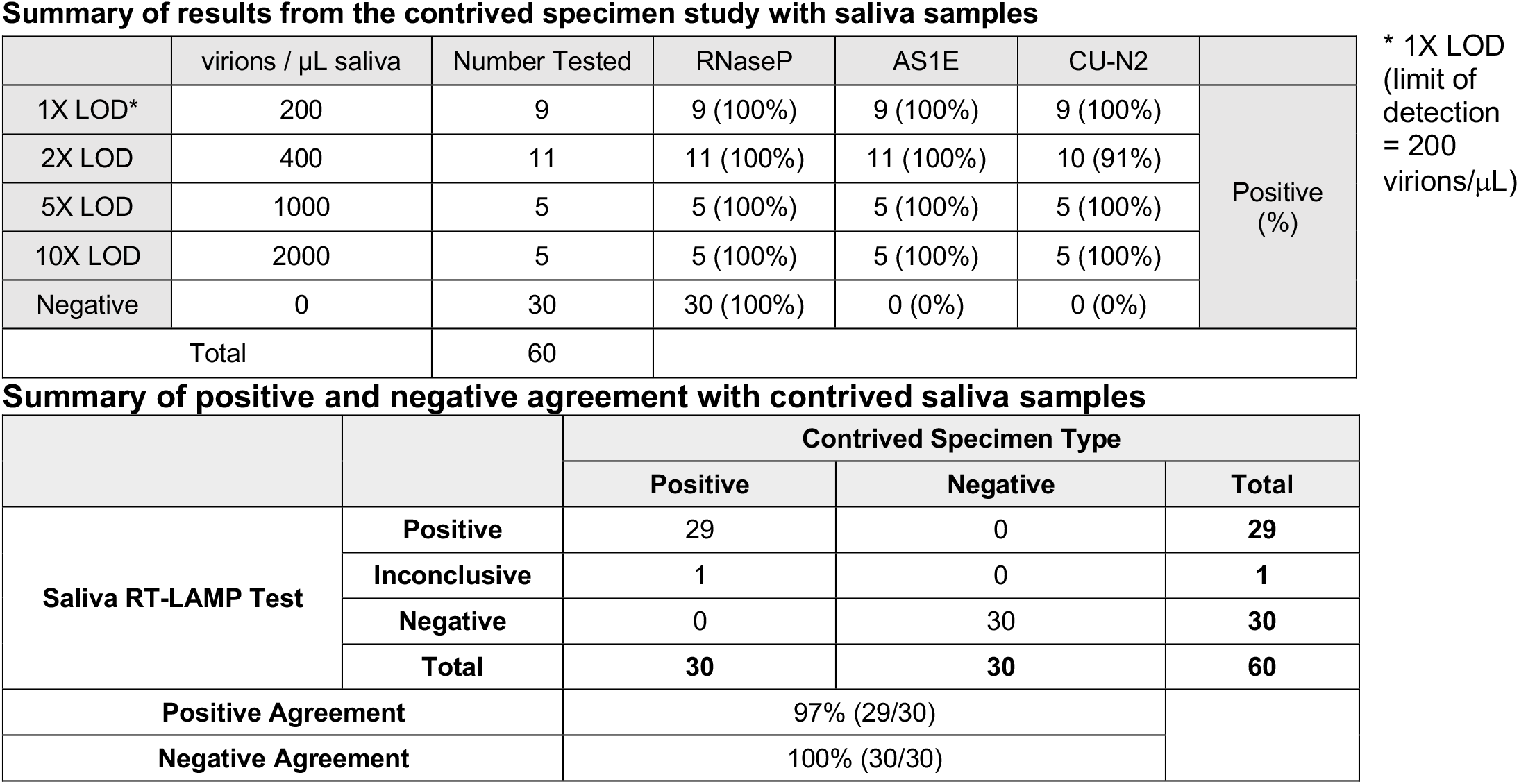
Blinded sample evaluation. Plain saliva, or saliva spiked with heat-inactivated SARS-CoV-2 virions at different concentrations, was heated at 95°C for 10 minutes. Samples were then analyzed using RT-LAMP by a researcher that did not know the true state of each sample. **Experiments in figure)** For each sample, three reactions were performed as indicated by each triplet of tubes. By looking at the patterns of yellow and pink results in each triplet, samples were scored according to the table below. The true status and observed result of each sample are listed to the right (P = Positive, N = Negative, I = Inconclusive). A white box on the triplet is shown if the sample contained SARS-CoV-2. One sample resulted in inconclusive test result. This sample did have SARS-CoV-2 spiked into it, but one of the SARS-CoV-2 primer sets failed to produce a signal (CU-N2). This failed reaction is still pink (negative) even though the tube has 2xLOD virus. 1X LOD = 200 virions/μL. Summary statistics for this experiment are provided in second and third tables below.

**Supplemental Figure S5:**
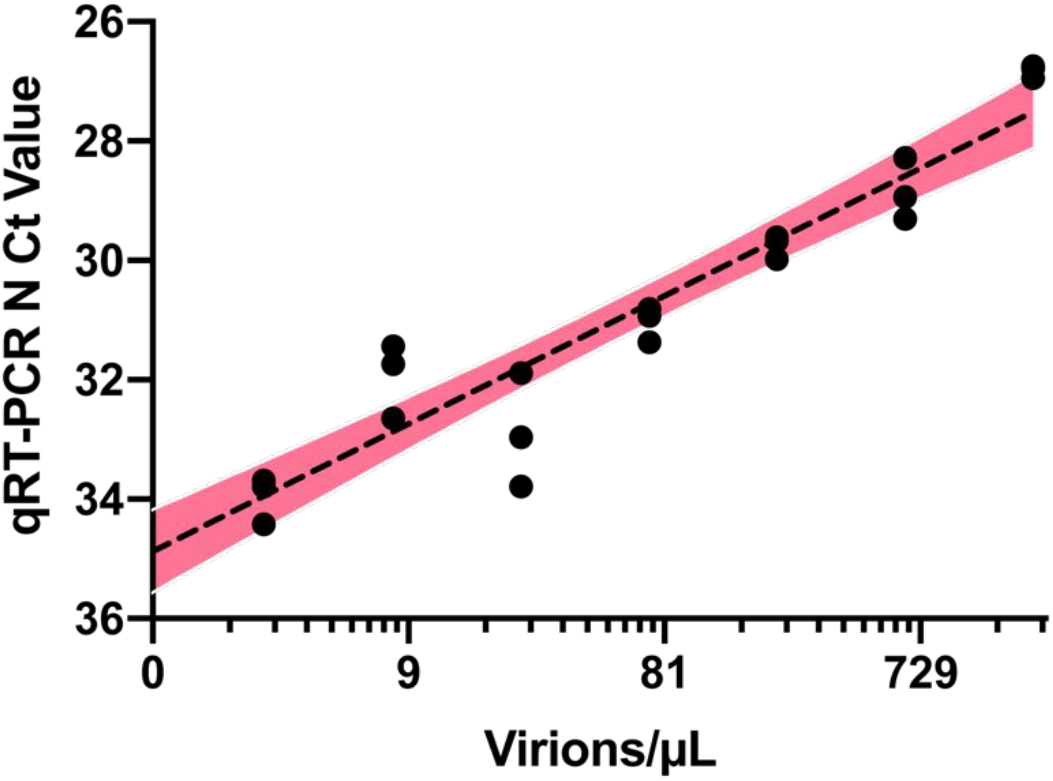
Quantitative RT-PCR standard curve used to determine the Ct value to virion/μL calculation. 10,000 copies/µL of heat deactivated SARS-CoV-2 virus was spiked into negative saliva specimens from 6 different volunteers and incubated for 30 minutes at 95°C. Samples were diluted to indicated concentrations using heat-treated saliva without SARS-CoV-2 addition from the same individual. The standard curve for the primer set targeting SARS-CoV-2 N gene is generated from the linear regression analysis and is illustrated with 95% confidence interval.

**Supplemental Figure S6.**
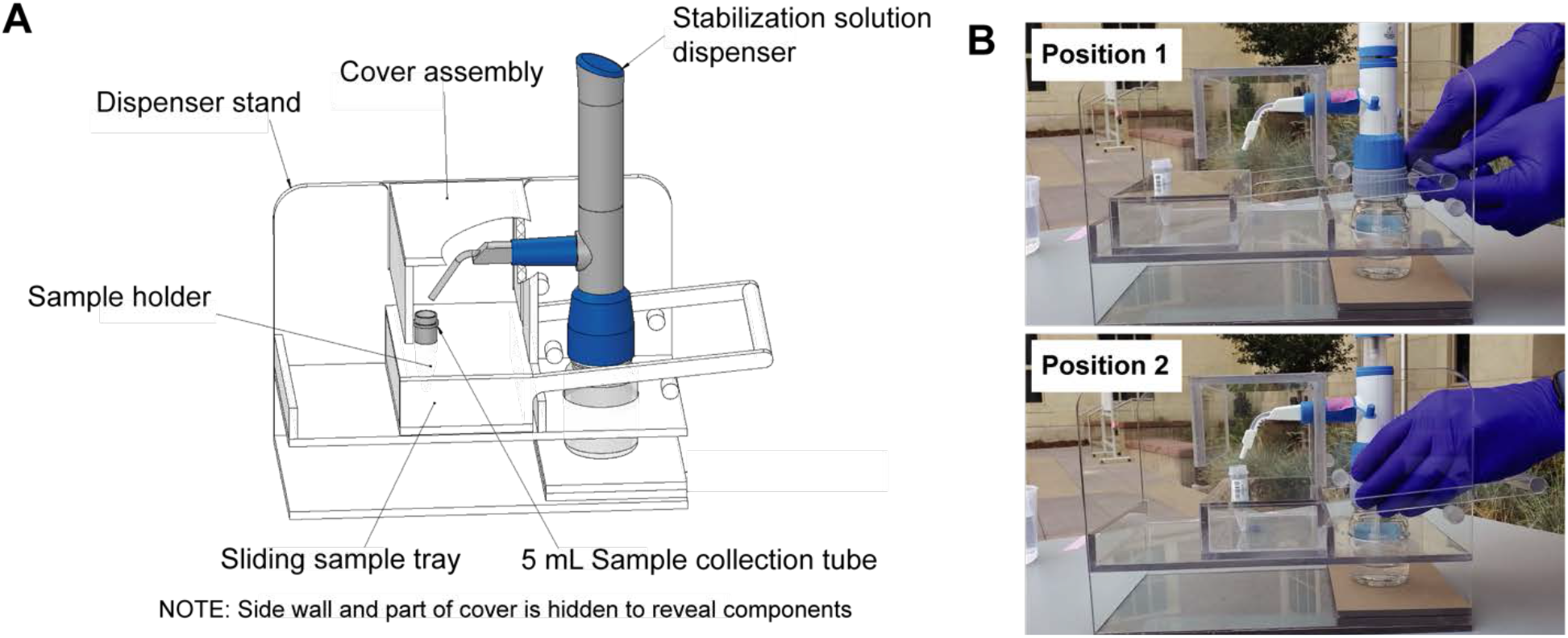
Diagrams of the saliva stabilization solution dispensing apparatus. **A)** CAD model of dispensing apparatus showing components. Custom solution dispensing apparatus fabricated from machined and solvent welded .236in polycarbonate (Tuffak). Polycarbonate is chosen for visibility, strength, and ability to withstand cleaning solvents such as ethanol. This device prevents the need for staff to directly handle uncapped and potentially infectious sample prior to inactivation, limits splash and aerosol exposure risk, and prevents cross-contamination of samples during solution addition step. **B)** Diagram illustrates the operation of the dispensing apparatus. **Position 1:** Tray is extended towards the testing participant and sample tube is seated in tray. Bottle containing stabilization solution and assembled with bottle top dispenser is seated in back section of apparatus. Staff moves tray towards themselves by gently pulling on handle until the tray is seated against back wall of the apparatus. **Position 2:** Sample tray is positioned against the back wall of the apparatus. This brings the sample tube and dispenser nozzle into a set orientation underneath the removable cover assembly. Sample collector uses bottle top dispenser to add 1mL of stabilization solution to sample then pushes sample tray back to the testing participant.

**Supplemental Figure S7.**
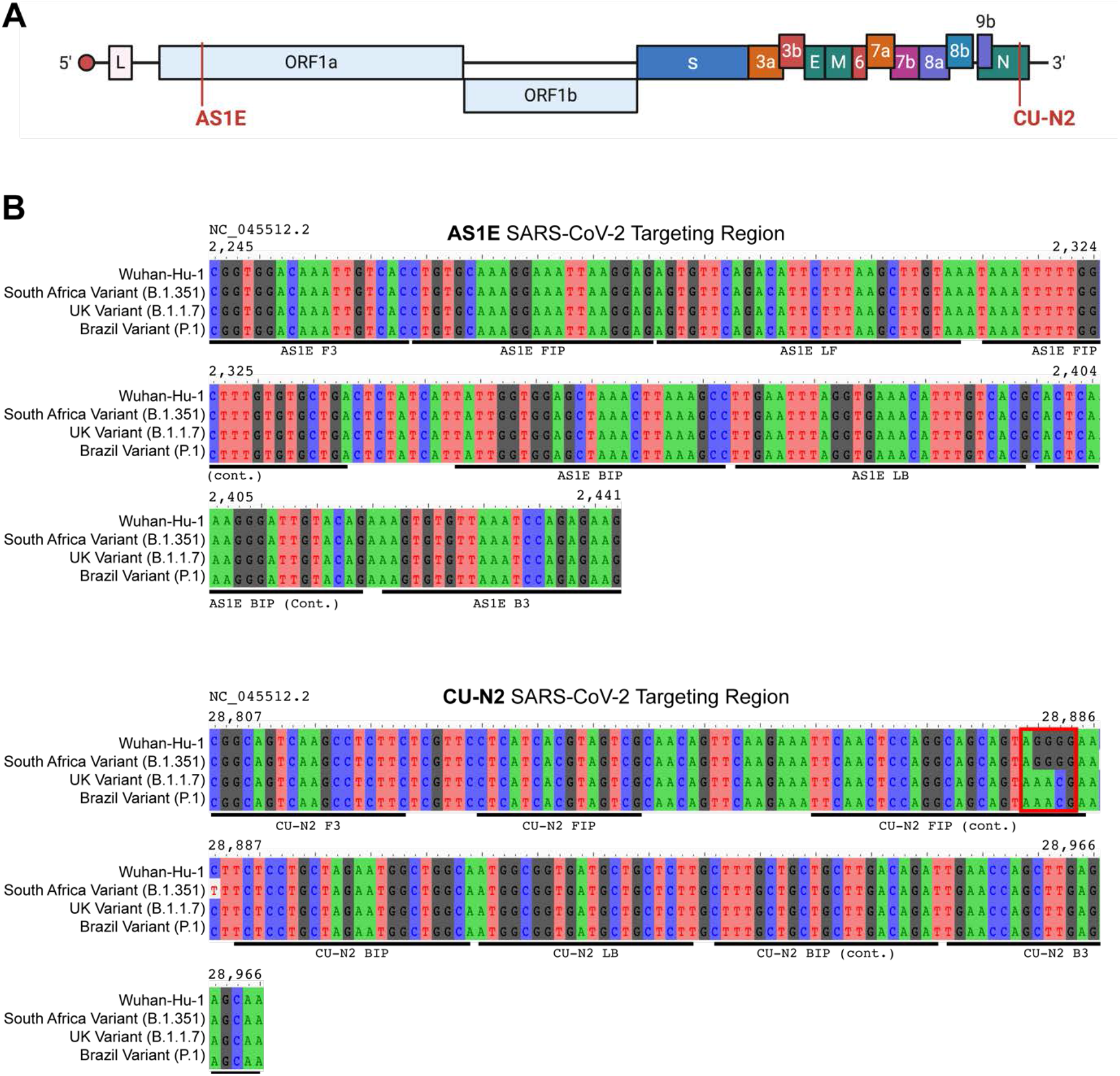
Saliva TwoStep primers will detect most or all currently circulating viral variants of concern. **A)** Genome map of SARS-CoV-2 with the regions targeted RT-LAMP primers highlighted in red. SARS-CoV-2 genome map is adapted from BioRender. **B)** Sequence alignments of regions of the key SARS-CoV-2 genome variants targeted by RT-LAMP primer sets AS1E and CU-N2. Binding regions of each individual primer set component is highlighted in underlying horizontal bars. The SARS-CoV-2 genome region targeted by AS1E primer set is conserved among all variants. For CU-N2, the red box highlights region of sequence variation that might render CU-N2 primer set less effective to identify the UK and Brazil variants. The coordinate of the genome sequence is based on the SARS-CoV-2 reference genome (NCBI NC_045512.2). The SARS-CoV-2 variant representative genomes are downloaded from GISAID (South Africa Variant B.1.351: hCoV-19/South Africa/KRISP-EC-K004572/2020; UK Variant B.1.1.7: hCoV-19/England/MILK-9E2FE0/2020; Brazil Variant P.1: hCoV-19/USA/VA-DCLS-2185/2020).

## Supplemental Text S1: Troubleshooting RT-LAMP False-Positives

See also Thi et al (Thi et al., 2020) for additional, excellent troubleshooting advice

## Controlling acidic and variable human saliva samples

We found that the biggest obstacle to implementing the colorimetric RT-LAMP assay is the variability in the reaction pH condition. First, false-positive signal can result from saliva samples that are naturally acidic. We spent significant time addressing this issue and ultimately found that all samples must be rendered basic as described in the article in order to set the correct threshold for specificity in the test. This was achieved through a titration series of sodium hydroxide in the saliva stabilization solution to find the optimal concentration that would ensure all RT-LAMP reactions start pink and are still capable of turning yellow if amplification occurs. However, note that other components in the saliva stabilization solution (EDTA, pH=8.0, and TCEP-HCl) have also played a role in establishing the optimal pH. Additional saliva stabilization solution optimization might be needed if other forms of these components are used. Second, the colorimetric RT-LAMP reaction relies on phenol red to detect the pH change during the amplification. Any additional buffering agents, such as tris-acetate or tris-borate that are commonly present in laboratory reagents, should be avoided to prevent potential false negative signals, as these buffering agents tend to inhibit the pH change.

## Controlling reaction acidification by carbonic acid

A second issue that has to be carefully controlled is the exposure of reaction components to the surrounding environment. When carbon dioxide from the atmosphere dissolves in water, it creates carbonic acid, which if present in high enough quantities can trigger the phenol red to turn yellow regardless of reaction state. Control measures should be implemented in three ways: First, we advise preparing the reaction mix (RT-LAMP master mix, primers, and water) right before sample loading. This is to prevent background amplification as well as the acidification of the reaction mix due to exposure to air. For this reason, we advise against the use of 96-well plates.

Additionally, dry ice should be avoided or completely isolated from the reaction components during sample transportation, as the exposure to the excessive carbon dioxide could also lead to acidification of the reaction mix. Second, during the 30-minute 65°C incubation, it is essential to completely seal off the reaction tubes to prevent vaporization of the reaction mix, as well as the infiltration of the water vapor if a water bath is used. We have noticed that an incomplete seal could lead to false positive signals. Last, because RT-LAMP amplification is highly robust, the test is very sensitive to contamination (Davidi et al., 2020). Therefore, opening of reaction tubes after RT-LAMP has occurred should be strictly avoided as these tubes contain a large amount of target DNA. Alternatively, the NEB WarmStart LAMP 2X Master Mix with UDG (NEB M1804) can be used to eliminate DNA contamination. Through these results are not shown, we have verified the same limit of detection can be reach using this alternative master mix.

## Controlling laboratory-based contamination

(Please also see Davidi et al, 2020 (Davidi et al., 2020)) When carrying out the RT-LAMP SARS-CoV-2 screening test at scale, it is critical to assign isolated workstations, each containing their own set of laboratory equipment such as pipettes, centrifuges, vortexes, and cleaning supplies. This equipment should never move between stations and be regularly decontaminated using a detergent based cleaning solution such as 10% bleach or any other commercially available solution designed to eliminated nucleic acid contaminants. Additionally, special care should be taken by laboratory staff to regularly replace gloves if moving *backwards* from the following workstations:

### Workstation 1: Setting up master mixes

This workstation is dedicated to making and aliquoting the master mix containing the RT-LAMP enzymes, primers, and water. All reagents should be centrifuged and spun down after thawing. As mentioned above, master mix should be made and aliquoted shortly before addition of saliva (Workstation 2) to avoid carbon dioxide solubilization due to atmospheric exposure.

### Workstation 2: Adding saliva samples to aliquoted RT-LAMP reaction tubes

This workstation is dedicated to handling the processed saliva samples (see **Supplemental Text S2** for advice on collecting and processing saliva). Once saliva samples are added to the aliquoted RT-LAMP reaction tubes, care should be taken to ensure that an appropriate seal is established (e.g. dome cap strips) to minimize airflow during reaction incubation. This workstation should include two micropipettes capable of pipetting a volume of 2 µl. One pipette can be used for saliva samples, while the other pipette should be used exclusively for pipetting any RNA controls (e.g. *in vitro* transcribed SARS-CoV-2 RNA).

### Workstation 3: RT-LAMP reaction incubation and results reporting

This workstation contains the heating element (e.g. heat block, thermal cycler) where RT-LAMP reactions are incubated. This Workstation has the highest risk of contamination, since the RT-LAMP reaction products will be in high abundance and can themselves serve as a template in subsequent reactions. When reactions are removed from the heating element, they should immediately be analyzed, results recorded, and then reaction tubes should be disposed in a container with a lid. *Never carry completed reaction tubes to any other part of the lab. Never open completed reaction tubes for any reason*. Any laboratory technician that has entered Workstation 3 should dispose of their gloves before returning to any other part of the lab.

## Supplemental Text S2: Recommended procedures for sample collection

When deploying Saliva TwoStep RT-LAMP screening test, it is important to set up saliva sample collection sites that allow large numbers of participants to move through the sample collection process quickly and smoothly. In addition, it is important to exercise extra precautions to avoid sample cross contamination as well as the exposure of the sample or stabilization solution. With this in mind, we have designed and optimized a saliva sample collection workflow that utilizes a customized stabilization dispensing apparatus (**Supplemental Figure S6**):

1. At the designated sample collection site, the screening test participants retrieve a 5 mL screw cap tube (MTC Bio #C2530) and collect passive drool into the 5mL tube until liquid saliva reaches the 1mL graduation mark. Bubbles do not contribute towards the 1mL volume.
2. After collecting saliva, the testing participants submerge capped 5 mL collection tube in a 250mL beaker with 70% ethanol to decontaminate the surface. They then remove the tube, carefully uncap it, and place it in the slot of the dispensing apparatus sample tray while in **Position 1** (**Supplemental Figure S6B**).
3. Staff then move the sample tray towards themselves by gently pulling on sample tray handle until it is seated against the back wall and the sample tube is underneath the cover assembly and centered under the dispenser nozzle (**Supplemental Figure S6B Position 2**). This partially enclosed space limits the potential risk of splashes and aerosols during this solution addition step.
4. Staff then use the bottle top dispenser (Fisher Scientific #13681527) to gently add 1mL of stabilization solution into sample tube (for an approximate 1:1 ratio of sample to solution). The apparatus holds the dispenser nozzle and sample tube in a fixed orientation to prevent cross contamination during this step.
5. Staff slide the sample tray containing the collection tube back towards the participant. The participant re-approaches to cap their sample with the screw-top lid, shakes it vigorously for 5-10 seconds to mix, cleans the surface with a wipe or by dunking in disinfectant, and places it on ice.
6. Staff then heat-inactivates the sample on-site by incubating it in a 95°C water bath or heat block (heat block preferred for minimizing spill risk) for 10 minutes.
7. Before the next testing participant approaches, staff sprays the sample tray with disinfectant.
8. Staff subsequently stores the inactivated sample on ice in a cooler and then transports it to Saliva TwoStep RT-LAMP testing area (see **Supplemental Text S1**).

### Biosafety note

Staff involved in saliva collection should wear all appropriate PPE including a fit-tested N95 mask. Regular surface decontamination is performed with 70% ethanol or bleach in the case of spills. Heating elements and cords are secured and situated away from foot traffic. Collection takes place outdoors for ventilation purposes whenever possible. Subjects maintain 10-foot distance from each other while unmasked and producing samples. Hand sanitizer is provided before and after collection procedure. Protocols were approved by the Institutional Biosafety Committee.

### Collection site material list

Staff set up a table with the following supplies: (1) 250mL plastic beaker with 70% ethanol, (1) 250mL plastic beaker with 10% bleach, (2) spray bottles with 70% ethanol, (1) dispenser apparatus (custom polycarbonate device) with bottle top reagent dispenser (Fisher) and 100mL glass bottle with stabilization solution, (1) ice bucket with ice, (1) water bath with tube rack and temperature probe, (1) digital timer, (1) cooler with ice. Additional items include paper towels, trash receptacles, spill kits, power cords, hand sanitizer, and additional PPE, ethanol, and bleach. Quantities can be scaled up as needed.

